# SARS-CoV-2 booster vaccination rescues attenuated IgG1 memory B cell response in primary antibody deficiency patients

**DOI:** 10.1101/2022.07.14.22276948

**Authors:** Frank J. Lin, Alexa Michelle Altman Doss, Hannah G. Davis-Adams, Lucas J. Adams, Christopher H. Hanson, Laura A. VanBlargan, Chieh-Yu Liang, Rita. E. Chen, Jennifer Marie Monroy, H. James Wedner, Anthony Kulczycki, Tarisa L. Mantia, Caitlin C. O’Shaughnessy, Saravanan Raju, Fang R. Zhao, Elise Rizzi, Christopher J. Rigell, Tiffany Biason Dy, Andrew L. Kau, Zhen Ren, Jackson Turner, Jane A. O’Halloran, Rachel M. Presti, Daved H. Fremont, Peggy L. Kendall, Ali H. Ellebedy, Philip A. Mudd, Michael S. Diamond, Ofer Zimmerman, Brian J. Laidlaw

**Author notes:** Correspondence should be addressed to Brian Laidlaw and Ofer Zimmerman. These authors contributed equally (Frank J. Lin, Alexa Michelle Altman Doss).

## Abstract

SARS-CoV-2 vaccines have proven effective in eliciting an immune response capable of providing protective immunity in healthy individuals. However, whether SARS-CoV-2 vaccination induces a long-lived immune response in immunocompromised individuals is poorly understood. Primary antibody deficiency (PAD) syndromes are among the most common immunodeficiency disorders in adults and are characterized by an impaired ability to mount robust antibody responses following infection or vaccination. Here, we present data from a prospective study in which we analyzed the B and T cell response in PAD patients following SARS-COV-2 vaccination. Unexpectedly, individuals with PAD syndromes mounted a SARS-CoV-2 specific B and CD4^+^ T cell response that was comparable in magnitude to healthy individuals. Many individuals with PAD syndromes displayed reduced IgG1^+^ and CD11c^+^ memory B cell responses following the primary vaccination series. However, the IgG1 class-switching defect was largely rescued following mRNA booster vaccination. Boosting also elicited an increase in the SARS-CoV-2-specific B and T cell response and the development of Omicron-specific memory B cells in COVID-19-naïve PAD patients. Together, these data indicate that SARS-CoV-2 vaccines elicit memory B and T cells in PAD patients that may contribute to long-term protective immunity.

## INTRODUCTION

Severe acute respiratory syndrome coronavirus 2 (SARS-CoV-2) is the causative agent of COVID-19 and has infected more than 500 million individuals resulting in over 6 million deaths as of June 2022. The mRNA-based Pfizer-BioNTech (BNT162b2) and Moderna (mRNA-1273) and the vector-based Johnson & Johnson (Ad26.COV2.S) SARS-CoV-2 vaccines are approved for use in the United States and have demonstrated efficacy in preventing symptomatic and asymptomatic infection (*1*–*7*). Although SARS-CoV-2-specific antibody titers wane over time, a durable cellular immune response is detectable for at least 6 months following completion of the primary vaccination series (*8*). The administration of an mRNA booster vaccine dose leads to a rapid increase in antibody titers and enables robust neutralization of viral variants, including Omicron (B.1.1.529), which can evade immunity elicited by the primary vaccination (*9*–*12*).

Individuals with medical conditions that compromise their ability to mount immune responses, such as primary and secondary immunodeficiencies, are at increased risk for severe illness and death following SARS-CoV-2 infection (*13, 14*). Patients with primary and secondary immunodeficiencies also have an impaired SARS-CoV-2-specific antibody response following a primary vaccination series (*15*–*24*). Moderately or severely immunosuppressed patients are recommended by the Centers for Disease Control and Prevention (CDC) to receive a third dose as part of their primary series against SARS-CoV-2 and a fourth dose at least 3 months following the completion of the primary vaccination series. Administration of booster doses leads to an increased SARS-CoV-2-specific antibody response in immunocompromised individuals (*17, 24*).

Primary antibody deficiency (PAD) syndromes are the most common symptomatic primary immunodeficiency in adults and are characterized by an impaired ability to mount an antibody response following infection or vaccination (*25*). The etiology of PAD syndromes is unknown in most patients, with only 25-35% of cases explained by inborn errors of immunity (*26*–*30*). Individuals with PAD syndromes are at increased risk of recurrent and severe infections, autoimmunity, allergic disease, and cancer (*25*). Most individuals with PAD syndromes receive intravenous or subcutaneous immunoglobulin replacement therapy every 1 to 4 weeks to reduce the frequency and severity of infections (*31*). However, immunoglobulin replacement therapy consists of immunoglobulin donated up to one year earlier and is unlikely to contain high titers of neutralizing antibodies specific for the strain of SARS-CoV-2 that is dominant at the time of administration (*24, 32, 33*).

We previously found that COVID-19-naïve individuals with PAD syndromes had a reduced SARS-CoV-2-specific antibody response following vaccination relative to healthy donors (*24*). The administration of a booster vaccine dose increased the antibody response and led to the development of antibodies with an enhanced ability to neutralize the Delta (B.1.617.2) and Omicron variants (*24*). However, the total and neutralizing antibody titers markedly declined by day 90 post-boost suggesting that the endogenous antibody response may be insufficient to mediate long-term protective immunity in individuals with PAD syndromes (*24*). In this study, we performed a prospective analysis of the SARS-CoV-2-specific B and T cell response following SARS-CoV-2 primary and booster vaccination in PAD patients. Unexpectedly, we found that most individuals with PAD syndromes generated a memory B and T cell response that was comparable in magnitude to the response in healthy donors following the primary vaccination series. Administration of a booster dose led to a further enhancement in B and T cell responses, including the development of Omicron-specific B cells. This work provides important insight into the capacity of memory B and T cells to contribute to protective immunity in PAD patients.

## RESULTS

### Individuals with PAD syndromes display a similar memory B cell response to healthy controls following primary vaccination series

We assessed the SARS-CoV-2-specific B and CD4^+^ T cell response following vaccination in peripheral blood mononuclear cells (PBMCs) from a cohort of 30 individuals with PAD syndrome (n=20 common variable immunodeficiency (CVID), n=4 hypogammaglobulinemia, n=6 specific antibody deficiency) (**Fig. 1a, Table S1**). This cohort completed their primary vaccination series (n=19 Pfizer-BioNTech BNT162b2, n=8 Moderna mRNA-1273, n=3 J&J Ad26.COV2.S) with 9 of these individuals having a real time-polymerase chain reaction (RT-PCR)-confirmed history of prior COVID-19 infection that occurred 36 to 276 days prior to vaccination. 19 of these individuals subsequently received a booster vaccine dose (n=16 BTN162b2, n=3 mRNA-1273). PBMCs were obtained from these individuals at multiple time points following completion of the primary vaccine series or the booster vaccine dose. PBMCs also were obtained from a separate cohort of 11 COVID-19-naïve healthy donors following completion of the primary vaccination series (n=11 BNT162b2) (**Table S1**). We then used flow cytometry to assess the immune cell response in PBMCs following SARS-CoV-2 vaccination (**Fig. 1b, S1a, Table S2**). PAD patients had a reduced percentage of B and T cells relative to healthy donors (**Fig. S1b, c, Table S2**). However, there was no difference in the percentage of B cells that were IgD^lo^ or IgD^lo^CD27^+^ or in the ratio of CD4^+^ to CD8^+^ cells among the T cell population (**Fig. S1b, c, Table S2**). 29 out of 30 PAD patients had an absolute lymphocyte count within the normal range on their most recent complete blood count (**Table S3**).

**Figure 1.**
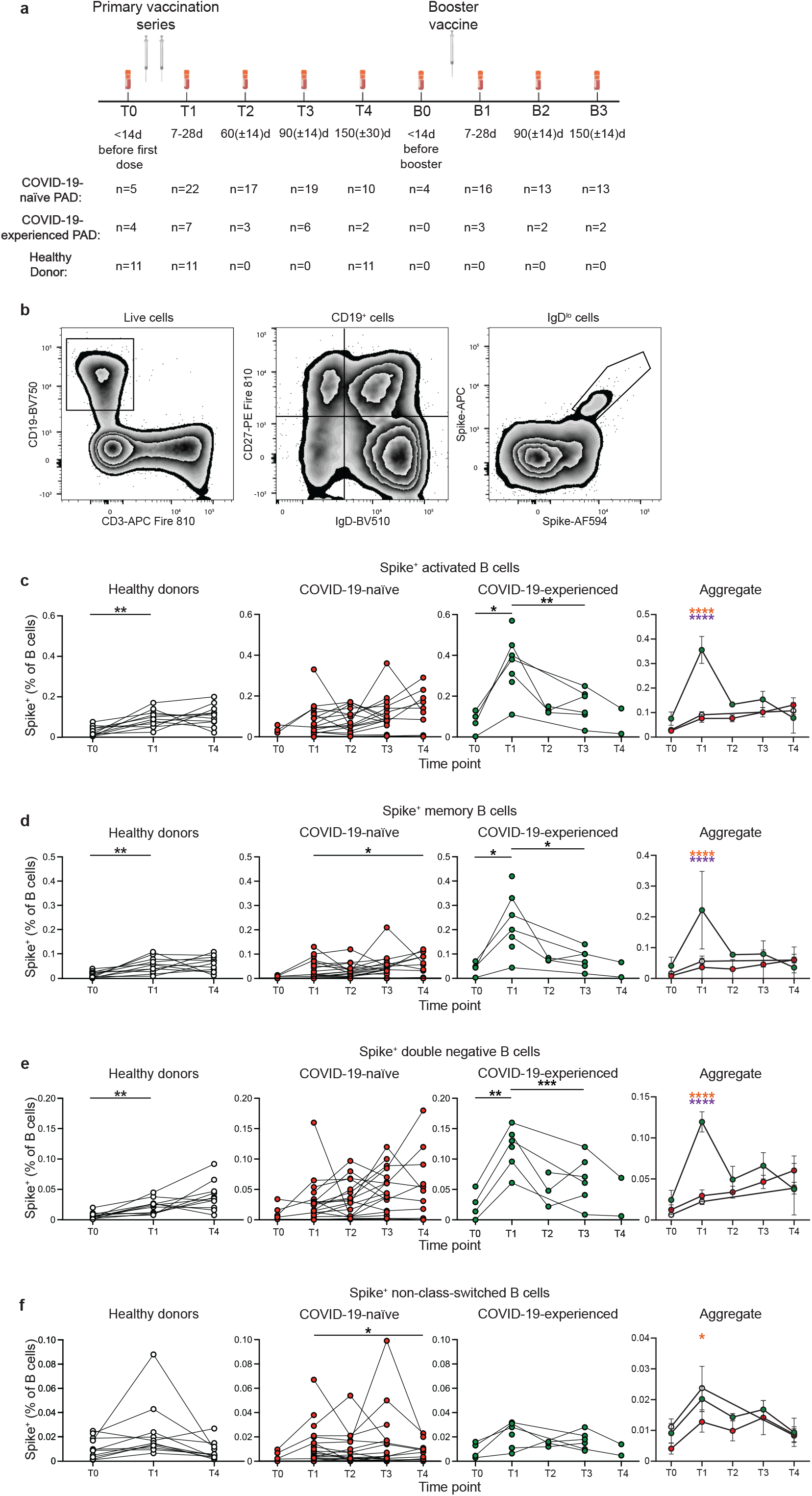
COVID-19-experienced PAD patients display elevated spike-specific B cell response following primary vaccination series. (**a**) Schematic of study design including time points in which PBMCs were obtained and number of samples per time point for each group. (**b**) Representative FACS plots of the gating strategy used to identify activated (IgD^lo^) Spike^+^ B cells. (**c**) Percentage of activated Spike^+^ cells amongst the B (Live CD19^+^ CD3^-^) cell population in the healthy donor (left, white), COVID-19-naïve PAD (middle left, red), and COVID-19-experienced PAD (middle right, green) cohorts. Aggregate of mean percentage of B cells that are Spike^+^ activated cells in all groups is shown on right. (**d**) Percentage of memory (IgD^lo^ CD20^+^ CD38^int-lo^ CD27^+^) Spike^+^ cells amongst the B cell population in the healthy donor (left, white), COVID-19-naïve PAD (middle, red), and COVID-19-experienced PAD (middle, green) cohorts. Aggregate of mean percentage of B cells that are Spike^+^ memory cells in all groups is shown on right. (**e**) Percentage of double negative (IgD^lo^ CD20^+^ CD38^int-lo^ CD27^-^) Spike^+^ cells amongst the B cell population in the healthy donor (left, white), COVID-19-naïve PAD (middle, red), and COVID-19-experienced PAD (middle, green) cohorts. Aggregate of mean percentage of B cells that are double negative Spike^+^ cells in all groups is shown on right. (**f**) Percentage of non-class-switched (IgD^+^ CD20^+^ CD38^int-lo^ CD27^+^) Spike^+^ cells amongst the B cell population in the healthy donor (left, white), COVID-19-naïve PAD (middle, red), and COVID-19-experienced PAD (middle, green) cohorts. Aggregate of mean percentage of B cells that are Spike^+^ non-class-switched cells in all groups is shown on right. Statistical analyses in c-f were performed using a mixed-effects model (for trends found between time points) or two-way ANOVA (for trends found between groups in the aggregate graphs) with Fisher’s least significant difference testing. Significance testing between time points was limited to comparisons relative to T1. On the aggregate graphs, error bars were displayed based on the standard error of the mean. Above the aggregate graphs, an orange asterisk indicates a comparison between the COVID-19-naïve and COVID-19-experienced groups, and a purple asterisk indicates a comparison between the COVID-19-experienced and healthy donor groups (*, *p* < 0.05; **, *p* < 0.01; ***, p<0.001; ****, p<0.001). See also Figure S2.

We then used His-tagged spike and RBD-binding probes to identify SARS-CoV-2-specific B cells (*34, 35*). We found that 25 of 29 (86%) PAD patients with available samples had a detectable spike and receptor binding domain (RBD)-specific IgD^lo^ B cell response following vaccination (**Fig. 1c, S2a**). Unexpectedly, we found that most COVID-19-naïve PAD patients displayed a comparable SARS-CoV-2-specific B cell response to COVID-19-healthy donors at all time points (**Fig. 1c, S2a**). The four PAD patients that did not respond to vaccination had a reduced percentage of B cells that were IgD^lo^ and IgD^lo^ CD27^+^ and had a reduced neutralizing antibody titer against the WA1/2020 and B.1.617.2 viruses following vaccination compared to responding PAD patients (**Fig. S1b, d, Table S2**). COVID-19-experienced individuals with PAD syndrome displayed a greater SARS-CoV-2 specific B cell response at day 7 to 28 following vaccination relative to both healthy donor and COVID-19 naïve PAD patients, before declining to a comparable level to the other 2 groups by day 60 (**Fig. 1c, S2a**). The SARS-CoV-2-specific response amongst the activated B cell (IgD^lo^ CD20^+^ CD38^int-lo^) population was divided between conventional (CD27^+^) and double negative (CD27^-^) memory B cells, with both populations displaying similar kinetics in COViD-19-naïve PAD patients and healthy donors (**Fig. 1d, e, S2b, c**). Double negative B cells accumulate in individuals with chronic infection or autoimmunity, but also are induced following vaccination in healthy individuals (*36, 37*). There was also a small population of SARS-CoV-2-specific B cells detected amongst the non-class-switched memory (IgD^+^ CD27^+^) B cell population following vaccination, with this population declining to baseline levels by day 150 in all groups (**Fig. 1f, S2d**). Together, these data indicate that most individuals with PAD syndromes can induce a comparable B cell response following SARS-CoV-2 vaccination to healthy donors, and that prior exposure to COVID-19 leads to a greater response upon vaccination in PAD patients.

### COVID-19-naïve individuals with PAD syndrome have a robust B cell response following booster vaccination

We next evaluated the B cell response following booster vaccination in individuals with PAD syndromes. Most COVID-19-naïve individuals with PAD syndromes mounted a SARS-CoV-2-specific B cell response at day 7-28 following booster vaccination, with an elevated percentage of cells present at day 150 following boosting compared to pre-boost levels (**Fig. 2a, S3a**). However, there was minimal increase in the SARS-CoV-2-specific B cell response in COVID-experienced PAD patients following boosting (**Fig. 2a-d, S3a-d**). The SARS-CoV-2-specific B cell response following boosting was largely composed of conventional memory and double negative B cells (**Fig. 2b-d, S3b-d**). While the percentage of SARS-CoV-2-specific memory B cells prior to booster vaccination did not correlate with the titer of neutralizing antibodies against the WA1/2020 and B.1.617.2 viruses, there was a correlation between the RBD-specific memory B cell response and the neutralizing antibody titer against B.1.1.529 (**Fig. 2e-g**). This suggests that SARS-CoV-2-specific memory B cells present in PAD individuals prior to booster vaccination can give rise to antibody-secreting cells capable of neutralizing viral variants.

**Figure 2.**
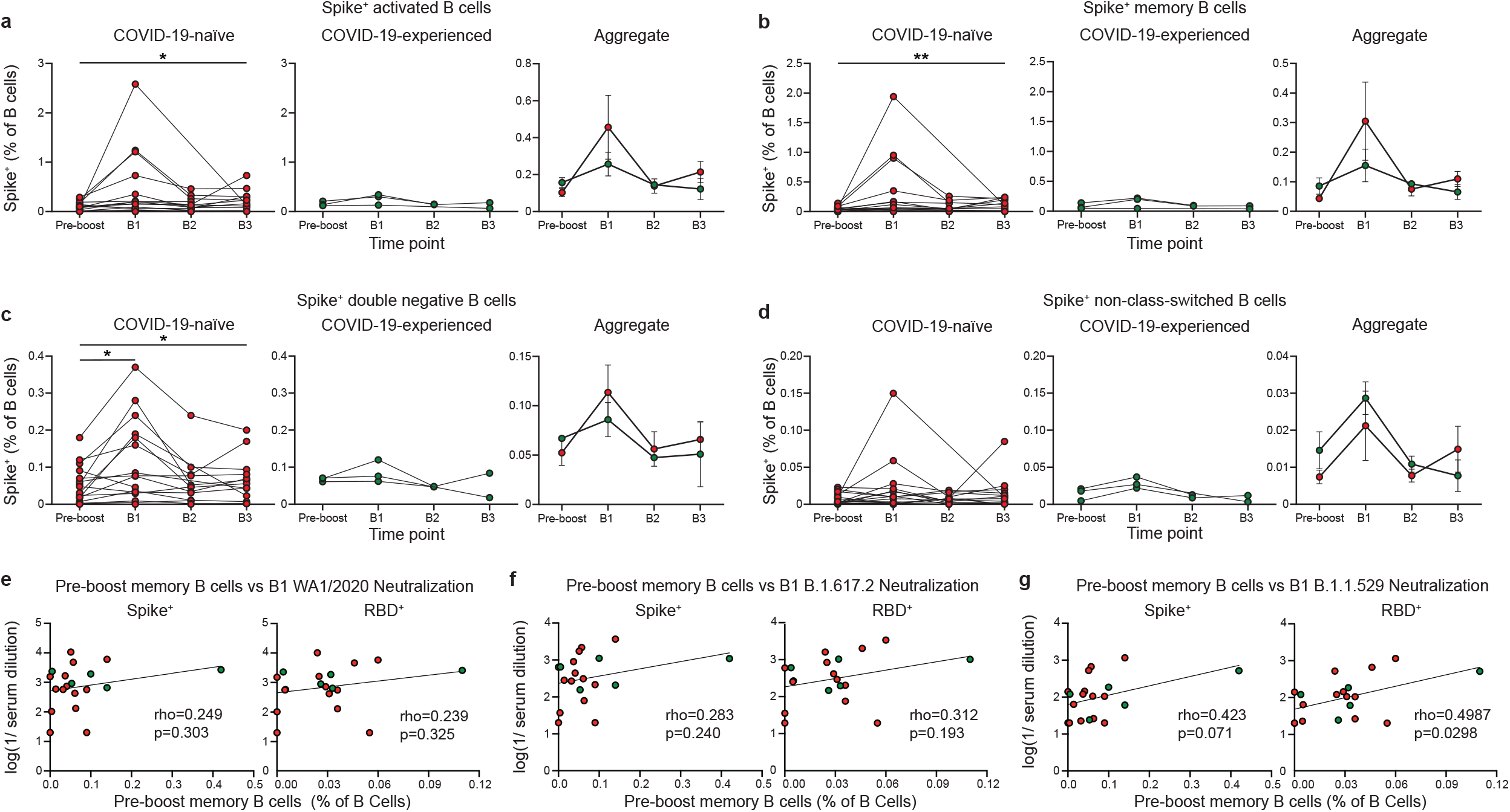
COVID-19-naive PAD patients display elevated spike-specific B cell response following booster vaccination. (**a**) Percentage of activated Spike^+^ cells amongst the B (Live CD19^+^ CD3^-^) cell population in the healthy donor (left, white), COVID-19-naïve PAD (middle left, red), and COVID-19-experienced PAD (middle right, green) cohorts. Aggregate of mean percentage of B cells that are Spike^+^ activated cells in all groups is shown on right. (**b**) Percentage of memory (IgD^lo^ CD20^+^ CD38^int-lo^ CD27^+^) Spike^+^ cells amongst the B cell population in the healthy donor (left, white), COVID-19-naïve PAD (middle, red), and COVID-19-experienced PAD (middle, green) cohorts. Aggregate of mean percentage of B cells that are Spike^+^ memory cells in all groups is shown on right. (c) Percentage of double negative (IgD^lo^ CD20^+^ CD38^int-lo^ CD27^-^) Spike^+^ cells amongst the B cell population in the healthy donor (left, white), COVID-19-naïve PAD (middle, red), and COVID-19-experienced PAD (middle, green) cohorts. Aggregate of mean percentage of B cells that are double negative Spike^+^ cells in all groups is shown on right. (**d**) Percentage of non-class-switched (IgD^+^ CD20^+^ CD38^int-lo^ CD27^+^) Spike^+^ cells amongst the B cell population in the healthy donor (left, white), COVID-19-naïve PAD (middle, red), and COVID-19-experienced PAD (middle, green) cohorts. Aggregate of mean percentage of B cells that are Spike^+^ non-class-switched cells in all groups is shown on right. (**e**) Correlation between percentage of B cells that are Spike^+^ (left) or RBD^+^ (right) memory cells prior to administration of booster vaccination and the serum neutralizing activity against WA1/2020 at B1. (**f**) Correlation between percentage of B cells that are Spike^+^ (left) or RBD^+^ (right) memory cells prior to administration of booster vaccination and the serum neutralizing activity against B.1.617.2 at B1. (**g**) Correlation between percentage of B cells that are Spike^+^ (left) or RBD^+^ (right) memory cells prior to administration of booster vaccination and the serum neutralizing activity against B.1.1.529 at B1. The pre-boost group consists of the last sample obtained from each patient prior to booster vaccination. Statistical analyses were performed using a mixed effects model (for trends found between time points) with Fisher’s least significant difference testing in a-d, or a Pearson rank correlation (with Pearson trend lines for visualization) in e-g. (*, *p* < 0.05; **, *p* < 0.01). Significance testing between time points was limited to comparisons relative to pre-boost. On the aggregate graphs, error bars were calculated based on the standard error of the mean. See also Figure S3.

### Defect in IgG1 class-switching in memory B cells from COVID-19-naïve individuals with PAD syndrome rescued following booster vaccination

We next evaluated the isotype specificity of the conventional memory B cell response following SARS-CoV-2 vaccination in 25 PAD patients that responded to vaccination (**Fig. 3a**). COVID-19-naïve individuals with PAD syndromes had a reduced percentage of IgG1^+^ memory B cells relative to healthy donors at day 7 to 28 post vaccination, with these individuals also displaying an elevated percentage of IgM^+^ SARS-CoV-2-specific memory B cells (**Fig. 3b, c, S4a, b**). However, booster vaccination led to an increase in the percentage of IgG1^+^ memory B cells in COVID-19-naïve PAD patients to levels comparable to the healthy donors post vaccination (**Fig. 3b, S4a**). Booster vaccination also led to a concomitant decrease in the percentage of IgM^+^ memory B cells (**Fig. 3c, S4b**). There was no significant difference in the percentage of IgA^+^, IgG2^+^ or IgG3^+^ memory B cells between healthy donors and COVID-19-naïve individuals (**Fig. 3d-f, S4c-e**). COVID-19-experienced individuals with PAD syndromes displayed a similar memory B cell isotype composition to the healthy donor cohort following the primary vaccination series (**Fig. 3, S4**). Together, these data indicate that the repeated exposure to SARS-CoV-2 through vaccination and/or infection can rescue the defect in IgG1-class switching seen in some individuals with PAD syndrome.

**Figure 3.**
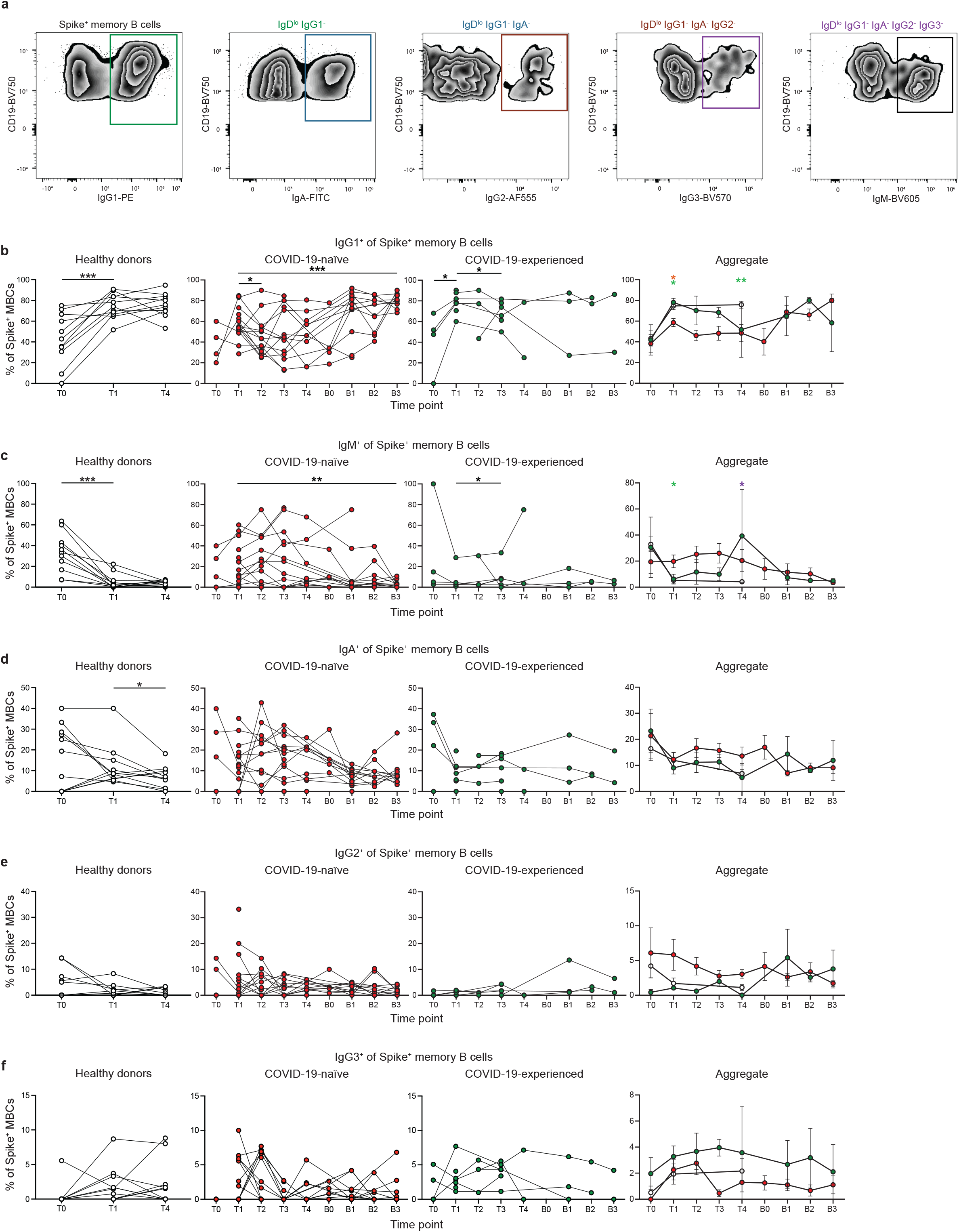
Spike-specific memory B cells from COVID-19-naive PAD patients display reduced IgG1 class switching following primary vaccination series. (**a**) Representative FACS plots of the gating strategy used to identify the isotype of Spike^+^ memory (IgD^lo^ CD20^+^ CD38^int-lo^ CD27^+^) B cells. (**b**) Percentage of Spike^+^ memory B cells that are IgG1^+^ in the healthy donor (left, white), COVID-19-naïve PAD (middle left, red), and COVID-19-experienced PAD (middle right, green) cohorts. Aggregate of mean percentage of IgG1^+^ cells in all groups is shown on right. (**c**) Percentage of Spike^+^ memory B cells that are IgM^+^ in the healthy donor (left, white), COVID-19-naïve PAD (middle left, red), and COVID-19-experienced PAD (middle right, green) cohorts. Aggregate of mean percentage of IgM^+^ cells in all groups is shown on right. (**d**) Percentage of Spike^+^ memory B cells that are IgA^+^ in the healthy donor (left, white), COVID-19-naïve PAD (middle left, red), and COVID-19-experienced PAD (middle right, green) cohorts. Aggregate of mean percentage of IgA^+^ cells in all groups is shown on right. (**e**) Percentage of Spike^+^ memory B cells that are IgG2^+^ in the healthy donor (left, white), COVID-19-naïve PAD (middle left, red), and COVID-19-experienced PAD (middle right, green) cohorts. Aggregate of mean percentage of IgG2^+^ cells in all groups is shown on right. (**f**) Percentage of Spike^+^ memory B cells that are IgG3^+^ in the healthy donor (left, white), COVID-19-naïve PAD (middle left, red), and COVID-19-experienced PAD (middle right, green) cohorts. Aggregate of mean percentage of IgG3^+^ cells in all groups is shown on right. Statistical analyses in b-d were performed using a mixed effects model (for trends found between time points) or two-way ANOVA (for trends found between groups in the aggregate graphs) with Fisher’s least significant difference testing. Significance testing between time points was limited to comparisons relative to T1. On the aggregate graphs, error bars were calculated based on the standard error of the mean. Above the aggregate graphs, a green asterisk indicates a comparison between the COVID-19-naïve and healthy donor groups, an orange asterisk indicates a comparison between the COVID-19-naïve and COVID-19-experienced groups, and a purple asterisk indicates a comparison between the COVID-19-experienced and healthy donor groups (*, *p* < 0.05; **, *p* < 0.01; ***, *p* < 0.001). See also Figure S4.

### Memory B cells from individuals with PAD syndrome display impaired CD11c expression following vaccination

We next assessed the phenotype of the SARS-CoV-2 specific conventional memory B cell response post vaccination (**Fig. 4a, b**). We observed a marked increase in the expression of CD11c on SARS-CoV-2-specific memory B cells at day 7 to 28 following vaccination in all groups, (**Fig. 4c, S5a**). CD11c expression is induced on B cells following antigen encounter and therefore distinguishes recently activated cells. (*38*). There was also an increase in the percentage of CD11c^+^ cells following boosting in the COVID-19-naïve PAD syndrome group (**Fig. 4c, S5a**). However, even following an mRNA booster dose, CD11c expression in COVID-19-naïve and experienced PAD patients was reduced relative to healthy donors at day 7 to 28 post vaccination, suggesting that there is impaired B cell activation in some individuals with PAD syndromes (*39*) (**Fig. 4c, S5a**). There was a significant correlation between the percentage of CD11c^+^ and IgG1^+^ Spike^+^ memory B cells at day 7 to 28 post vaccination in COVID-19-naive PAD patients (**Fig. 4d, S5b**). CD11c expression can be induced on B cells by B cell receptor (BCR) signaling (*38*). This suggests that the defect in IgG1 class switching seen in some PAD patients may be related to impaired BCR signaling. However, there was no correlation between the percentage of CD11c^+^ and IgG1^+^ CD27^+^ Spike^+^ memory B cells following boosting indicating that repeated antigen exposure can rescue the defect in IgG1 class-switching despite the reduction in B cell activation in many PAD patients (**Fig. 4d, S5b)**. The percentage of CD11c^+^ memory B cells decreased by day 90 post vaccination or boosting, with this decrease accompanied by a concomitant increase in the percentage of CXCR5^+^ memory B cells in al groups (**Fig. 4c, e, S5a, b**).

**Figure 4.**
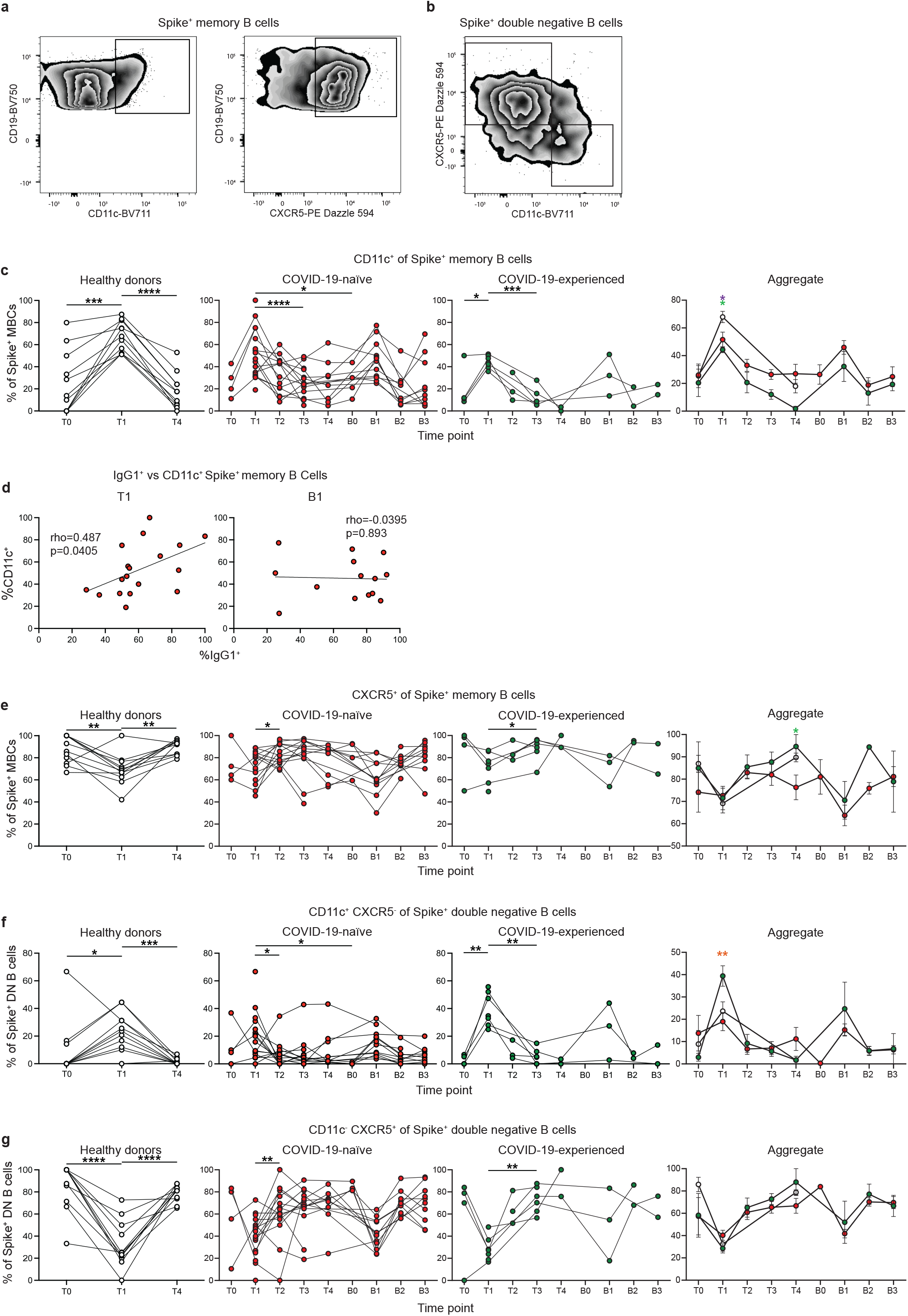
Spike-specific memory B cells from PAD patients display reduced CD11c expression class. (**a**) Representative FACS plots of the expression of CD11c and CXCR5 on Spike^+^ memory (IgD^lo^ CD20^+^ CD38^int-lo^ CD27^+^) B cells. (**b**) Representative FACS plots of the expression of CD11c and CXCR5 on Spike^+^ double negative (IgD^lo^ CD20^+^ CD38^int-lo^ CD27^-^) B cells. (**c**) Percentage of Spike^+^ memory B cells that are CD11c^+^ in the healthy donor (left, white), COVID-19-naïve PAD (middle left, red), and COVID-19-experienced PAD (middle right, green) cohorts. Aggregate of mean percentage of CD11c^+^ cells in all groups is shown on right. (**d**) Correlation between percentage of Spike^+^ memory B cells that are IgG1^+^ and CD11c^+^ at T1 (left) or B1 (right). Associations for **d** are calculated using Pearson rank correlation and are shown with Pearson trend lines for visualization. (**e**) Percentage Spike^+^ memory B cells that are CXCR5^+^ in the healthy donor (left, white), COVID-19-naïve PAD (middle left, red), and COVID-19-experienced PAD (middle right, green) cohorts. Aggregate of mean percentage of CXCR5^+^ cells in all groups is shown on right. (**f**) Percentage of Spike^+^ double negative B cells that are CD11c^+^ CXCR5^-^ in the healthy donor (left, white), COVID-19-naïve PAD (middle left, red), and COVID-19-experienced PAD (middle right, green) cohorts. Aggregate of mean percentage of are CD11c^+^ CXCR5^-^ cells in all groups is shown on right. (**g**) Percentage of Spike^+^ double negative B cells that are CD11c^-^ CXCR5^+^ in the healthy donor (left, white, COVID-19-naïve PAD (middle left, red), and COVID-19-experienced PAD (middle right, green) cohorts. Aggregate of mean percentage of are CD11c^-^ CXCR5^+^ cells in all groups is shown on right. Statistical analyses in c, e-g were performed using a mixed effects model (for trends found between time points) or two-way ANOVA (for trends found between groups in the aggregate graphs) with Fisher’s least significant difference testing. Significance testing between time points was limited to comparisons relative to T1. On the aggregate graphs, error bars were calculated based on the standard error of the mean. Above the aggregate graphs, a green asterisk indicates a comparison between the COVID-19-naïve and healthy donor groups, an orange asterisk indicates a comparison between the COVID-19-naïve and COVID-19-experienced groups, and a purple asterisk indicates a comparison between the COVID-19-experienced and healthy donor groups (*, *p* < 0.05; **, *p* < 0.01; ***, *p <* 0.001; ****, *p <* 0.0001). See also Figure S5.

We also determined the phenotype of the SARS-CoV-2 specific double negative B cell response following vaccination (*40*). Double negative B cells can be divided into subsets based on expression of CD11c and CXCR5 (*40*). There was an increased percentage of CD11c^+^ CXCR5^-^ (DN2) cells at day 7 to 28 post vaccination in all groups, with a higher increase in COVID-19 experienced PAD patients compared to COVID-19 naïve patients (**Fig. 4f, S5d**). A similar increase was observed in both groups of individuals with PAD syndromes following boosting (**Fig. 4f, S5d**). Conversely, there was a decrease in the percentage of CD11c^-^ CXCR5^+^ (DN1) cells at day 7 to 28 post vaccination in all groups, with this population returning to baseline at day 60 post vaccination (**Fig. 4g, S5e**). A similar decrease in the percentage of DN1 cells also was apparent following boosting in the PAD syndromes cohort (**Fig. 4f, S5d**). There was no clear difference in the phenotype of the SARS-CoV-2-specific double negative memory B cells between the COVID-19-naïve PAD individuals and the healthy donor group (**Fig. 4f, g, S5d, e**).

### Individuals with PAD syndrome display a robust SARS-CoV-2-specific CD4^+^ T cell response following vaccination

We next evaluated the T cell response following SARS-CoV-2 vaccination (**Fig. 5a**). SARS-CoV-2-specific CD4^+^ T cells were identified using a S_167-180_ tetramer, which binds an immunodominant SARS-CoV-2 spike epitope restricted by the HLA-DPB1*04:01 allele that is found at >40% frequency in many populations around the world (*41*). We also developed a S_816-830_ tetramer that is specific for an immunodominant region of the S-II portion of spike protein (*42, 43*). This region is highly conserved among coronaviruses and is also restricted to the HLA-DPB1*4:01 allele (*42*). 16 out of 30 individuals with PAD syndromes and 4 out of 11 healthy donor samples had detectable SARS-CoV-2-specific CD4^+^ T cell responses. COVID-naïve PAD patients displayed a similar percentage of SARS-CoV-2-specific CD4^+^ T cell response as healthy donors at day 7 to 28 post vaccination before contracting by day 150 (**Fig. 5b-d**). Boosting led to an increase in the SARS-CoV-2-specific CD4^+^ T cell response (**Fig. 5b-d**). COVID-experienced PAD patients had an increased percentage of SARS-CoV-2-specific CD4^+^ T cell prior to vaccination relative to the other groups, consistent with a pre-existing memory response (**Fig. 5b-d**). There was no correlation between the magnitude of the SARS-COV-2 specific CD4^+^ T cell response and the conventional memory B cell response following vaccination in PAD patients (**Fig. 5e**). However, there was a strong correlation following boosting suggesting that the pre-existing memory response in PAD patients can give rise to an enhanced B and T cell response following antigen re-encounter (**Fig. 5f**).

**Figure 5.**
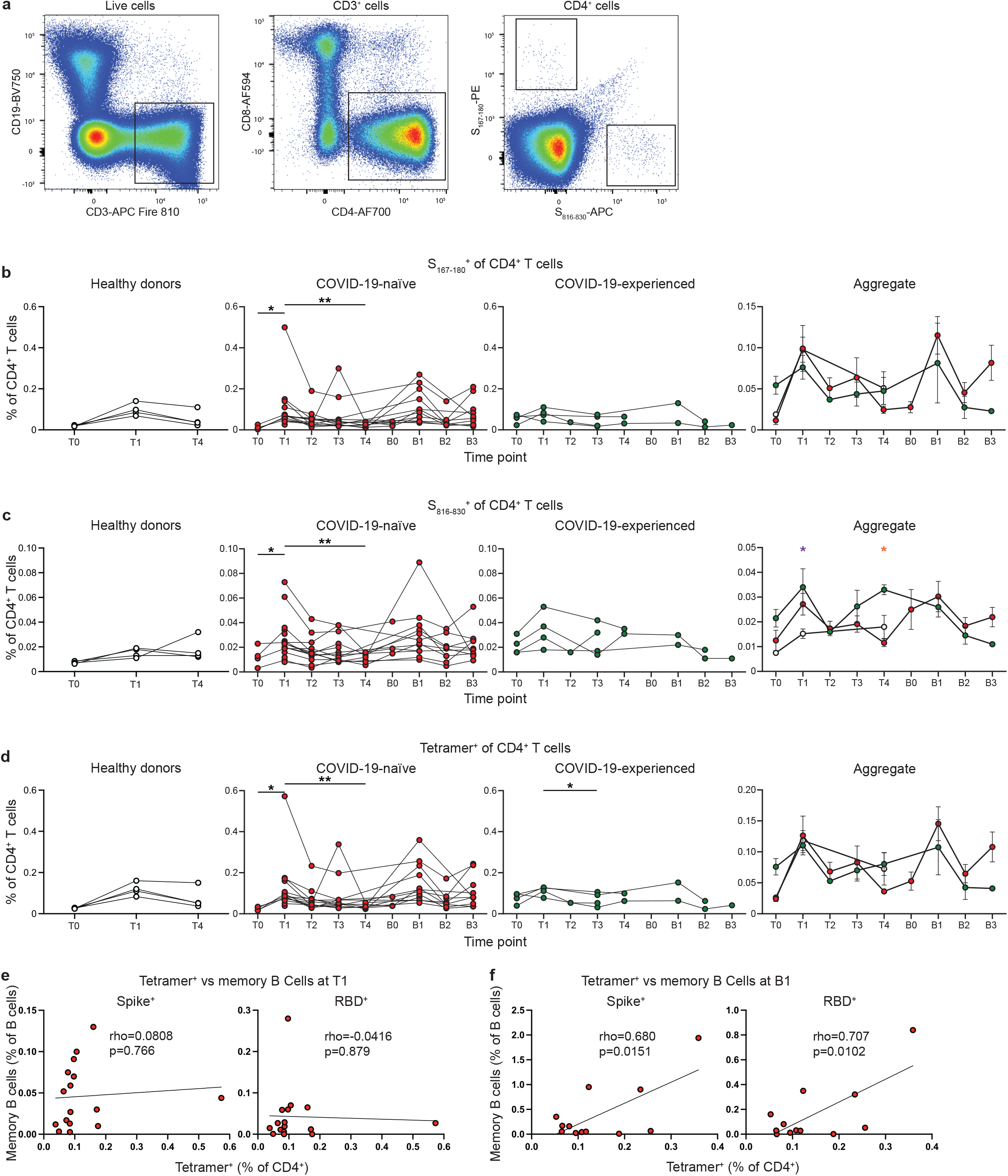
PAD patients display unimpaired SARS-CoV-2-specific CD4^+^ T cell response following vaccination. (**a**) Representative FACS plots of the gating strategy used to identify S_167-180_^+^ and S_816-830_^+^ CD4^+^ T cells. (**b**) Percentage of CD4^+^ (Live CD3^+^ CD19^-^ CD4^+^ CD8^-^) T cells that are S_167-180_^+^ in the healthy donor (left, white), COVID-19-naïve PAD (middle left, red), and COVID-19-experienced PAD (middle right, green) cohorts. Aggregate of mean percentage of S_167-180_^+^ cells in all groups is shown on right. (**c**) Percentage of CD4^+^ T cells that are S_816-830_^+^ in the healthy donor (left, white), COVID-19-naïve PAD (middle left, red), and COVID-19-experienced PAD (middle right, green) cohorts. Aggregate of mean percentage of S_816-830_^+^ cells in all groups is shown on right. (**d**) Combined percentage of CD4^+^ T cells that are tetramer^+^ (S_167-180_^+^ or S_816-830_^+^) in the healthy donor (left, white), COVID-19-naïve PAD (middle left, red), and COVID-19-experienced PAD (middle right, green) cohorts. Aggregate of mean percentage of tetramer^+^ cells in all groups is shown on right. (**e**) Correlation between percentage of B cells that are Spike^+^ (left) or RBD^+^ (right) memory (IgD^lo^ CD20^+^ CD38^int-lo^ CD27^+^) cells and the percentage of CD4^+^ T cells that are Tetramer^+^ at T1. (**f**) Correlation between percentage of B cells that are Spike^+^ (left) or RBD^+^ (right) memory cells and the percentage of CD4^+^ T cells that are Tetramer^+^ at B1. Associations for **e** and **f** are calculated using Pearson rank correlation and are shown with Pearson trend lines for visualization. Statistical analyses in b-d were performed using a mixed effects model (for trends found between time points) or two-way ANOVA (for trends found between groups in the aggregate graphs) with Fisher’s least significant difference testing. Significance testing between time points was limited to comparisons relative to T1. On the aggregate graphs, error bars were calculated based on the standard error of the mean. Above the aggregate graphs, an orange asterisk indicates a comparison between the COVID-19-naïve and COVID-19-experienced groups, and a purple asterisk indicates a comparison between the COVID-19-experienced and healthy donor groups (*, *p* < 0.05; **, *p* < 0.01).

### SARS-CoV-2-specific CD4^+^ T cells from PAD patients display a similar phenotype to cells from healthy donors

The phenotype of the SARS-CoV-2-specific CD4^+^ T cell response was next assessed (**Fig. 6a, b**). SARS-CoV-2-specific CD4^+^ T cells adopted an activated phenotype following vaccination and boosting in PAD patients, with this response characterized by increased cell surface expression of PD1, ICOS, and CD38 (**Fig. 6c-e**). Expression of these markers was similar between COVID-19-naïve individuals with PAD syndromes and heathy donors at day 7 to 28 following vaccination suggesting that there is no defect in CD4^+^ T cell activation in most individuals with PAD syndromes (**Fig. 6c-e**). We also did not detect a difference in HLA-DR expression between any of the groups (**Fig. 6f**). The phenotype of the CD4^+^ T cell response was further assessed by determining the percentage of SARS-CoV-2-specific central (CD45RO^+^CD27^+^CCR7^+^) and effector (CD45RO^+^CD27^+^CCR7^+^) memory CD4^+^ T cells (**Fig. 6g, h**). We found there was no difference in the composition of the memory T cell response at T4 between the groups, suggesting that there also not a defect in the development of memory CD4^+^ T cells in PAD patients (**Fig. 6g, h**). Together, these data indicate that patients with PAD syndromes exhibit a SARS-CoV-2-specific CD4^+^ T cell response that is similar in magnitude and quality to the T cell response in healthy donors following vaccination.

**Figure 6.**
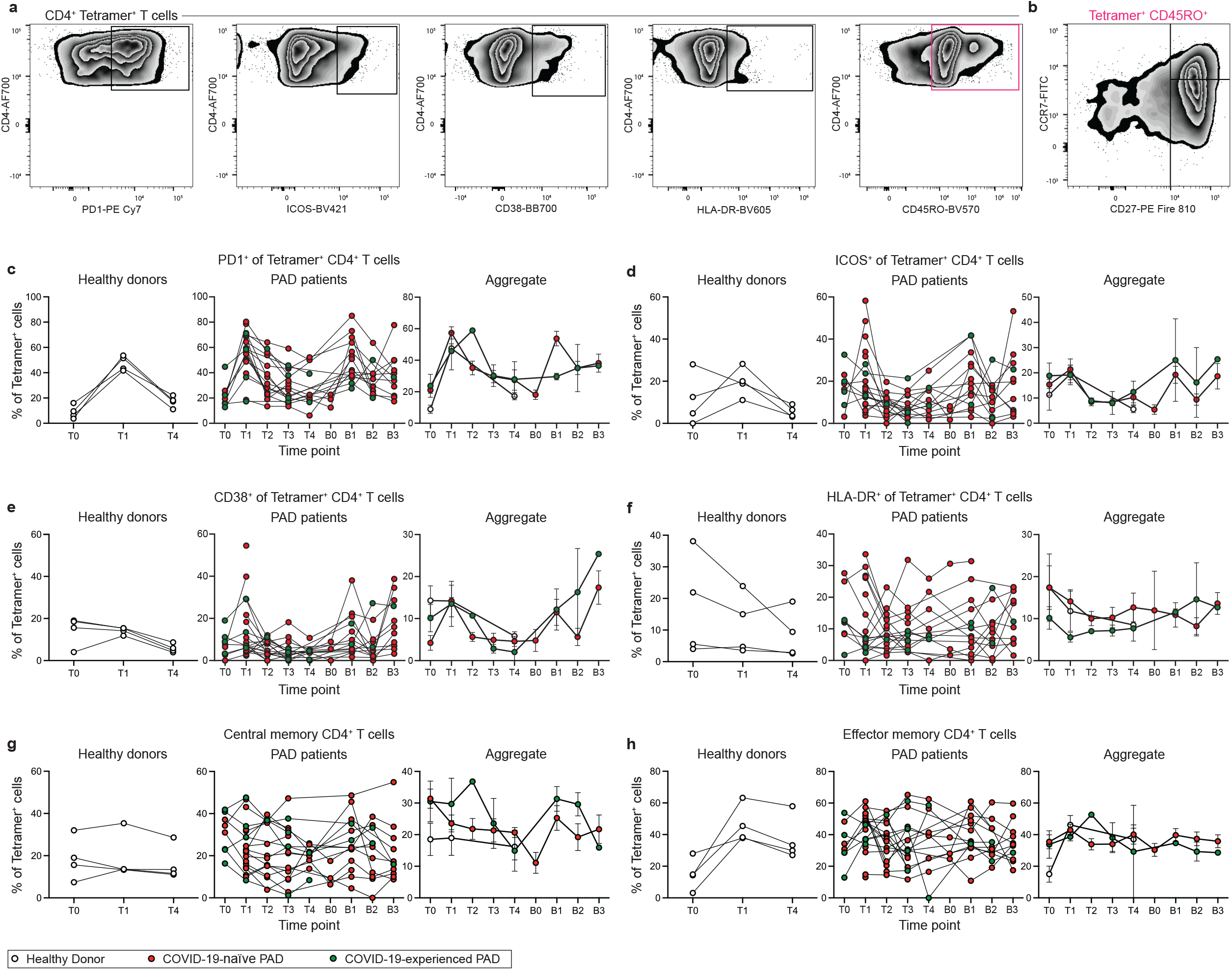
Phenotype of SARS-CoV-2-specific CD4^+^ T cell response following vaccination in PAD patients. (**a**) Representative FACS plots of the expression of PD1, ICOS, CD38, HLA-DR, and CD45RO on Tetramer^+^ CD4^+^ (Live CD3^+^ CD19^-^ CD4^+^ CD8^-^ S_167-180_^+^ or S_816-830_^+^) T cells. (**b**) Representative FACS plots of the expression of CD27 and CCR7 on CD45RO^+^ Tetramer^+^ CD4^+^ T cells. (**c**) Percentage of Tetramer^+^ CD4^+^ T cells that are PD1^+^ in the healthy donor (left, white), COVID-19-naïve PAD (middle, red), and COVID-19-experienced PAD (middle, green) cohorts. Aggregate of mean percentage of Tetramer^+^ T cells that are PD1^+^ in all groups is shown on right. (**d**) Percentage of Tetramer^+^ CD4^+^ T cells that are ICOS^+^ in the healthy donor (left, white), COVID-19-naïve PAD (middle, red), and COVID-19-experienced PAD (middle, green) cohorts. Aggregate of mean percentage of Tetramer^+^ T cells that are ICOS^+^ in all groups is shown on right. (**e**) Percentage of Tetramer^+^ CD4^+^ T cells that are CD38^+^ in the healthy donor (left, white), COVID-19-naïve PAD (middle, red), and COVID-19-experienced PAD (middle, green) cohorts. Aggregate of mean percentage of Tetramer^+^ T cells that are CD38^+^ in all groups is shown on right. (**f**) Percentage of Tetramer^+^ CD4^+^ T cells that are HLA-DR^+^ in the healthy donor (left, white), COVID-19-naïve PAD (middle, red), and COVID-19-experienced PAD (middle, green) cohorts. Aggregate of mean percentage of Tetramer^+^ T cells that are HLA-DR^+^ in all groups is shown on right. (**g**) Percentage of Tetramer^+^ T cells that are central memory (CD45RO^+^ CD27^+^ CCR7^+^) cells in the healthy donor (left, white), COVID-19-naïve PAD (middle, red), and COVID-19-experienced PAD (middle, green) cohorts. Aggregate of mean percentage of Tetramer^+^ T cells that are central memory cells in all groups is shown on right. (**h**) Percentage of Tetramer^+^ T cells that are effector memory (CD45RO^+^ CD27^+^ CCR7^-^) cells in the healthy donor (left, white), COVID-19-naïve PAD (middle, red), and COVID-19-experienced PAD (middle, green) cohorts. Aggregate of mean percentage of Tetramer^+^ T cells that are effector memory in all groups is shown on right. On the aggregate graphs, error bars were calculated based on the standard error of the mean.

### Booster vaccination induces Omicron-specific memory B cells in individuals with PAD syndrome

We assessed the Omicron-specific B cell response in individuals with PAD syndromes using a His-tagged protein specific for the spike protein of B.1.1.529 (BA.1) (**Fig. 7a**). We found that administration of a booster vaccine led to an increase in the percentage of Omicron specific memory B cells in COVID-19-naïve individuals (**Fig. 7b**). This increase was evident in both the conventional and double negative B cell populations (**Fig. 7c, d**). There was no increase in the percentage of Omicron-specific non-class switched B cells (**Fig. 7e**). The percentage of Omicron-specific B cell returned to pre-boost baseline level in most COVID-19-naïve individuals by day 90 post boost (**Fig. 7b-d**). However, patient 110 displayed a robust increase in their percentage of Omicron-specific B cell between B2 and B3, along with an increase in their SARS-CoV-2-specific T cell response and neutralizing antibody titers. The B2 and B3 time points correspond to the period between November 2021 and January 2022, when COVID-19 cases in the United States surged due to the emergence of the Omicron variant. Patient 102 was confirmed to have been re-infected between B2 and B3 and also displayed increased SARS-CoV-2-specific T cell, B cell, and neutralizing antibody responses. Of note, patient 102 and 110 reported no or very mild symptoms during this period.

**Figure 7.**
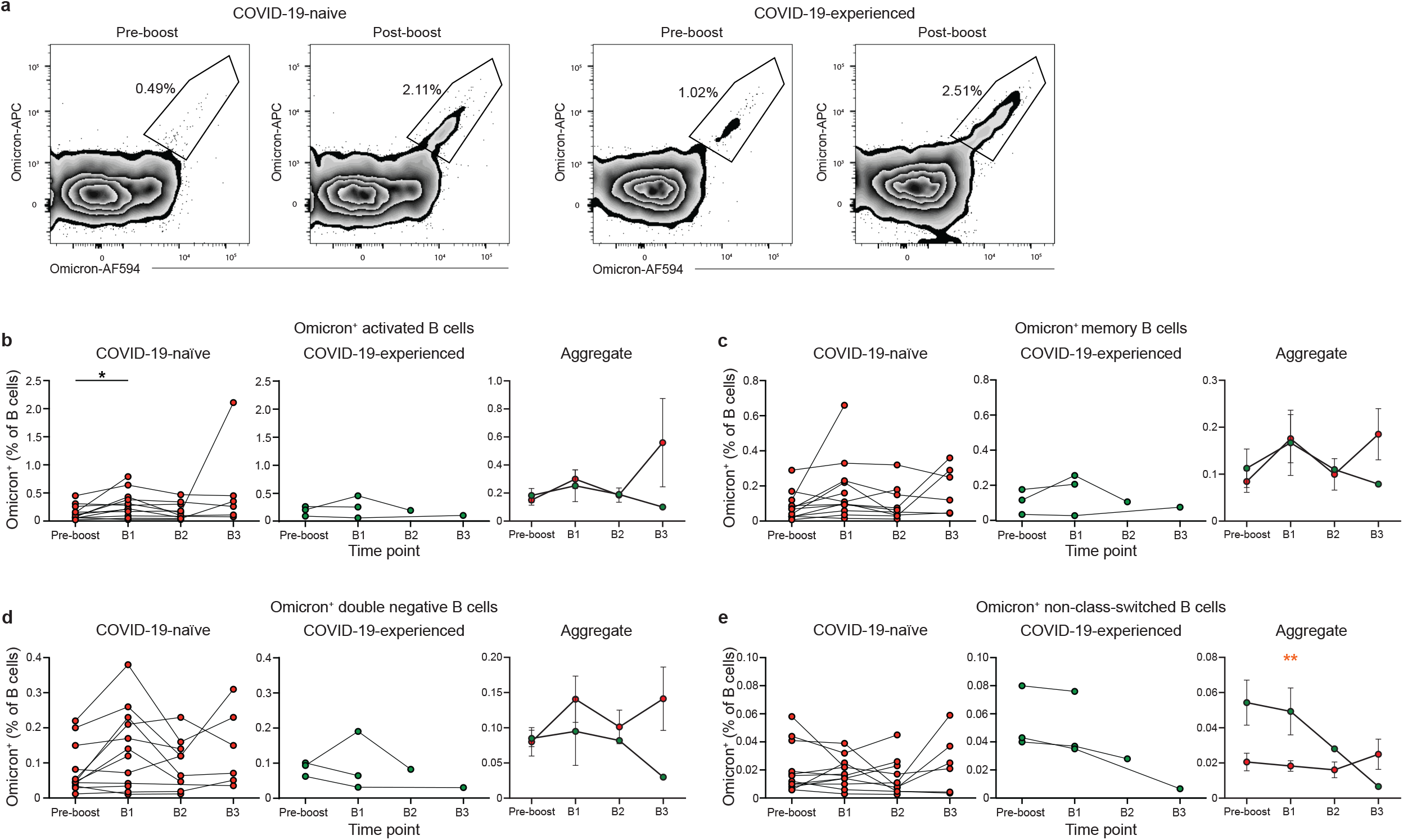
PAD patients display elevated Omicron-specific B cell response following booster vaccination. (**a**) Representative FACS plots of the percentage of Omicron-specific B cells among the memory (IgD^lo^ CD20^+^ CD38^int-lo^ CD27^+^) B cell population prior to and post booster vaccination in COVID-19-naïve (left) and COVID-19-experienced (right) individuals with PAD syndromes. (**b**) Percentage of activated Omicron^+^ cells amongst the B (Live CD19^+^ CD3^-^) cell population in the healthy donor (left, white), COVID-19-naïve PAD (middle left, red), and COVID-19-experienced PAD (middle right, green) cohorts. Aggregate of mean percentage of B cells that are Omicron^+^ activated cells in all groups is shown on right. (c) Percentage of memory Omicron^+^ cells amongst the B cell population in the healthy donor (left, white), COVID-19-naïve PAD (middle, red), and COVID-19-experienced PAD (middle, green) cohorts. Aggregate of mean percentage of B cells that are Omicron^+^ memory cells in all groups is shown on right. (**d**) Percentage of double negative Omicron^+^ cells amongst the B cell population in the healthy donor (left, white), COVID-19-naïve PAD (middle, red), and COVID-19-experienced PAD (middle, green) cohorts. Aggregate of mean percentage of B cells that are double negative Omicron^+^ cells in all groups is shown on right. (**e**) Percentage of non-class-switched (IgD^+^ CD20^+^ CD38^int-lo^ CD27^+^) Omicron^+^ cells amongst the B cell population in the healthy donor (left, white), COVID-19-naïve PAD (middle, red), and COVID-19-experienced PAD (middle, green) cohorts. Aggregate of mean percentage of B cells that are Omicron^+^ non-class-switched cells in all groups is shown on right. Statistical analyses in b-e were performed using a mixed effects model (for trends found between time points) or two-way ANOVA (for trends found between groups in the aggregate graphs) with Fisher’s least significant difference testing. Significance testing between time points was limited to comparisons relative to pre-boost. On the aggregate graphs, error bars were calculated based on the standard error of the mean. Above the aggregate graphs, an orange asterisk indicates a comparison between the COVID-19-naïve and COVID-19-experienced groups (*, *p* < 0.05; **, *p* < 0.01).

## DISCUSSION

In this study, we demonstrated that SARS-CoV-2 vaccination can induce a long-lived memory B and CD4^+^ T cell response in individuals with PAD syndromes that is comparable to the response seen in healthy donors. Furthermore, we found that the SARS-CoV-2-specific memory B cell response correlated with neutralizing antibody titers against the Omicron variant following boosting. Only 4 out of 29 patients did not develop a spike-specific memory B cell response following vaccination with these individuals also having lower levels of memory B cells and activated B cells compared to the other patients in the cohort. These results suggest that memory B and T cells can promote long-term protective immunity in individuals with PAD syndromes despite their impaired ability to mount optimal and sustained antibody responses following infection and vaccination.

Despite the normal total memory B cell response in PAD patients in our cohort, SARS-CoV-2-specific memory B cells from COVID-19-naïve PAD patients displayed reduced IgG1 class-switching following the primary vaccination series. These cells also displayed impaired CD11c expression, with a positive correlation seen between the percentage of spike-specific CD11c^+^ and IgG1^+^ cells following vaccination. CD11c expression is regulated by BCR stimulation, with CD11c^+^ B cells displaying increased expression of genes involved in B cell activation and antigen presentation (*38*). This suggests that the defect in IgG1 class-switching in some PAD patients may be due to impaired BCR signaling and/or reduced T cell help. IgG1 class switching is rescued in PAD patients following booster vaccination. mRNA booster vaccination also led to an increase in Omicron-specific B cells. Together, these data indicate that administration of mRNA booster vaccination doses may have durable benefits in addition to the short-term increase in total SARS-CoV-2-specific B cells and antibody titers. Many PAD patients in this cohort who historically had poor immune responses to bacterial and other protein antigens (e.g., Streptococcus pneumoniae polysaccharides, tetanus toxoid, and diphtheria toxin) as part of their initial immune workup responded to mRNA vaccines (**Table S4**). The basis of this difference remains unclear, although it could be due to the unique adjuvant properties of the lipid nanoparticles or in vitro-synthesized mRNA (*44*).

Our data also suggest that administration of additional booster vaccines may not lead to an additional short-term increase in SARS-CoV-2-specific B cells. COVID-19 experienced individuals with PAD syndrome had a robust response to the primary vaccination series but displayed no further increase in SARS-CoV-2-specific B cells after administration of a booster vaccine. These individuals also did not display any further increase in their percentage of IgG1^+^ memory B cells or in the antibody avidity (*24*). This finding is consistent with other studies showing that COVID-19-experienced healthy donors do not display a further increase in their SARS-CoV-2-specific memory B cell response following boosting (*45*). This does not indicate that additional booster vaccines may not promote additional evolution of the memory B cell repertoire in individuals with PAD syndromes that is independent of cell number (*46*).

While the B cell response after completion of the primary vaccination series was assessed in other work, these studies did not detect a SARS-CoV-2-specfic memory B cell response above baseline and concluded that vaccination of individuals with PAD syndromes primarily results in a double negative memory B cell response (*47, 48*). We find that SARS-CoV-2 vaccination induces both a conventional and double negative memory B cell response in most PAD patients that is comparable to healthy donors, and that this response is maintained for at least 150 days after completion of the primary vaccination series. The disparity in results between these studies may be due to a difference in sensitivity in the probes used, as the previous study detected very few cells that bound to spike probes even in healthy donors. Heterogeneity existing within individuals classified as having PAD syndromes may also contribute to difference seen between these studies.

T cells also have an important role in mediating protective immunity against SARS-CoV-2 (*44*). Assessment of the SARS-CoV-2-specific T cell response typically involves stimulation of PBMCs with peptides spanning the SARS-CoV-2 spike protein and assaying for activation induced markers (AIM) or intracellular cytokines. Previous studies reported a reduced percentage of interferon gamma-producing T cells following *ex vivo* stimulation in some vaccinated individuals with PAD syndromes (*15, 23, 48*). We stained PBMCs with tetramers against both S_167-180_ and S_816-830_ tetramers, which bind to immunodominant peptides restricted by the HLA-DPB1*04:01 allele. This allowed us to directly detect SARS-CoV-2 specific CD4^+^ T cells without requiring additional stimulation. The SARS-CoV-2-specific CD4^+^ T cell response was comparable in magnitude and phenotype to between COVID-naïve PAD patients and healthy donors that generated a detectable response. Booster vaccination led to a marked increase in the SARS-CoV-2-specfic CD4^+^ T cell response. These results suggest that the SARS-CoV-2-specific CD4^+^ T cells may contribute to the immune response upon re-infection. However, they do not exclude the possibility that SARS-CoV-2-specfiic CD4^+^ T cells in some individuals with PAD syndromes may exhibit impaired cytokine production. Further work is needed to assess the magnitude and phenotype of the SARS-CoV-2-specific CD8^+^ T cell response in PAD patients.

Individuals that are moderately to severely immunocompromised have an elevated risk of severe COVID-19 illness and death and are recommended to receive a third dose of mRNA-based vaccine as part of their primary series. This recommendation is supported by findings from our group and others that immunocompromised individuals have a reduced SARS-CoV-2-specific antibody titer following the initial two-dose vaccination series relative to healthy controls (*15, 23, 24, 47, 48*). Booster vaccination increased the SARS-CoV-2 specific antibody titer and led to the development of Omicron-specific neutralizing antibodies (*24*). However, the increase in neutralizing antibodies titers displayed following booster vaccination was short-lived and returned to the pre-boost baseline within 90 days (*24*). Our finding show that a booster dose increases the level and enriches the repertoire of spike-specific memory B cells in PAD patients and might have a long-term benefit beyond the short-lived increase in anti-spike and anti-RBD antibody titers.

Most individuals with PAD syndromes receive immunoglobulin replacement therapy. Immunoglobulin replacement products administered between May 2021-January 2022 had low levels of SARS-CoV-2-specific antibody titers with low neutralization activity against ancestral strains (*24*). While the titer of anti-spike and anti-RBD in immunoglobulin replacement products increased over time, the long lag of 9-12 months between collection and production of IVIG and SCIG commercial products may make most available commercial immunoglobulin replacement products less effective against current circulating Omicron variants (*33*). Although many individuals with PAD might be eligible for long-acting combination monoclonal antibody prophylaxis (e.g., Evusheld [AZD7442]) against COVID-19, recent studies showed substantial (∼176-fold) losses in potency against some lineages of Omicron virus (e.g., BA.1.1) (*49*). Therefore, immunization of individuals with PAD syndromes with mRNA vaccines that include a booster may be the most effective way to induce a protective immune response against SARS-CoV-2 and its variants.

### Limitations to the study

One limitation is that not all patients elected to receive a booster vaccination resulting in lower number of samples in the post-booster time points. This lack of samples is particularly apparent in the COVID-experienced group, which included only three individuals that received a booster vaccination, limiting the conclusions that can be drawn from this data. Another limitation is that the design of this study precluded the collection of a pre-vaccination blood draw from all individuals. Finally, it is important to note that PAD syndromes are a heterogeneous group of diseases with our cohort including individuals with CVID, hypogammaglobulinemia, and specific antibody deficiency. While we did not observe a clear difference in the immune response between these subgroups, there could be heterogeneity between different PAD subgroups that necessitate different vaccination and boosting approaches.

## MATERIALS AND METHODS

### Primary antibody deficiency syndromes cohort

The study was approved by the Institutional Review Board of Washington University School of Medicine (Approval # 202104138). Patients were identified by a medical record search for PAD syndromes, and their records were reviewed to confirm their diagnosis and verify they met the inclusion criteria. COVID-19 vaccination status was reviewed, and subjects were contacted if they were within the vaccination window or not yet immunized. Inclusion criteria included males and females over 18 years of age, health care provider-documented PAD syndromes including common variable immunodeficiency (CVID), specific-antibody deficiency, or hypogammaglobulinemia, and the ability to give informed consent. Entry criteria also included receipt of a SARS-CoV-2 vaccine within 14 days of enrollment, receipt of the second dose of mRNA vaccine (Moderna mRNA-1273 or Pfizer BioNTech BNT162b2) within 28 days of the first visit, or receipt of one dose of adenoviral-vector vaccine (J&J 3Ad26.COV2.S) within 35 days of initial visit. Exclusion criteria included participation in an investigational study of SARS-CoV-2 vaccines within the past year, history of HIV infection, an active cancer diagnosis, treatment with immunosuppressive medications, history of hematologic malignancy, history of anti-CD20 monoclonal antibody therapy, receipt of live-attenuated vaccine within 30 days or any inactivated vaccine within 14 days of SARS-CoV-2 vaccination, blood or blood product donation within 30 days prior to study vaccination, and planned blood donation at any time during or 30 days after the duration of subject study participation.

In total, 469 charts were reviewed, and 160 subjects were contacted. A total of 30 adults (27 females, 3 males) with PAD syndromes met eligibility requirements and agreed to enroll in the study (see Table S1); we note a sex-bias in the enrollees from our PAD cohort, which is not typical for the disease itself. Ages ranged from 20 to 82, with an average age of 48.4 years old. Twenty PAD patients had CVID, six had specific antibody deficiency, and four had hypogammaglobulinemia. Twenty-seven of these subjects had received immunoglobulin replacement therapy before and during the study period from nine different products. Nineteen subjects received the BNT162b2, eight received mRNA-1273, and three received Ad26.COV2.S vaccines. Of the 30 subjects, nine were diagnosed with a prior SARS-CoV-2 infection with a positive nasal swab RT-PCR test, and one received treatment with an anti-SARS-CoV-2 monoclonal antibody (bamlanivimab) 90 days prior to study enrollment (**Table S1**).

All subjects had one mandatory post-vaccine blood sample collection with optional pre-vaccine and follow-up visits at days 60, 90, and 150 (±14 days) after vaccination. The optional pre-vaccination blood sample was collected up to 14 days before receiving vaccine. For subjects who received a two-dose series of mRNA vaccines, the first post-vaccination blood collection occurred 7 to 28 days after the second dose. For subjects receiving the Ad26.COV2.S single-dose vaccine, the first post-vaccination blood sample was collected 21 to 35 days after immunization. Since the study was non-interventional, patients were informed if they mounted an immune response to the vaccine, but the decision to receive a booster was made between the patient and their physician. Subjects who opted for boosting provided a blood sample up to 14 days prior to receiving the booster dose, unless the subject previously provided a sample within 2 weeks as part of the optional post-vaccine assessments. Subjects returned for an additional sample 7 to 28 days after receiving the booster (range 7-27 days, median 17 days, mean 17 days. One patient had her post-booster sample drawn at day 35), with a second and third post-booster visit and sample collection occurring at 90 ±14 days and 150 ±14 days.

### Healthy donor cohort

Immunocompetent healthy donor volunteer blood samples were obtained as previously described (*50*). The healthy donor study was approved by the Institutional Review Board of Washington University School of Medicine (Approval # 202012081).

### Quantification and statistical analysis

Statistical significance was determined using Prism Version 9 (GraphPad). Statistical analysis was determined by one-way ANOVA, unpaired t-test, mixed model analysis, or two-way ANOVA with Fisher’s least significant difference testing. Associations were calculated using Pearson rank correlation and are shown with Pearson trend lines for visualization.

## Data Availability

All data reported in this paper will be shared upon request. Any additional information required to reanalyze the data reported in this paper is available from the corresponding authors upon request.

## ACKNOWLEDGEMENTS

This study was supported by grants and contracts from NIH: DP2AI169978 and K22AI153015 [all to B.J.L.], R01 AI157155, U01 AI151810, and 75N93019C00051 [all to M.S.D.], U01AI141990, U01AI150747, HHSN272201400006C, HHSN272201400008C, and 75N93019C00051 [all to A.H.E.], and R01 DK084242 [to P.L.K.]. The study also was supported by a VA Merit Award BX002882 [to P.L.K]. This study utilized samples obtained from the Washington University School of Medicine COVID-19 biorepository, which is supported by the NIH/National Center for Advancing Translational Sciences (UL1 TR002345). The WU368 studies were reviewed and approved by the Washington University Institutional Review Board (approval no. 308 202012081). The authors thank Likui Yang and the Bursky Center for Human Immunology Tetramer Core for their help generating tetramer reagents. The authors also express their gratitude to all study participants.

## AUTHOR CONTRIBUTIONS

F.J.L. performed experiments, analyzed the data, and prepared the manuscript.

A.L.T.D. enrolled subjects, collected demographic and clinical data, processed PBMC samples, performed experiments, and analyzed data.

H.G.D.A., F.R.Z. and E.R. processed PBMC samples.

L.J.A. and D.H.F. generated crucial reagents.

C.H.H. performed experiments and analyzed the data.

C.L., L.A.V, and R.E.C designed and performed antibody neutralization experiments and analyzed data.

J.M.M and C.J.R collected demographic and clinical data, enrolled patients, and provided patient care.

H.J.W., And.K, T.B.D, Ant.K. and Z.R. provided patient care.

T.L.M. wrote the study protocol, managed IRB compliance, enrolled patients, and processed patient samples.

C.C.O collected patient demographic and clinical data, enrolled patients, and processed samples.

S.R. planned experiments and analyzed data.

A.H.E and J.T contributed samples from the healthy donor cohort.

J.A.O and R.M.P wrote and maintained the Institutional Review Board protocol, recruited, and phlebotomized participants and coordinated sample collection of healthy donors.

P.L.K. supervised the project.

P.A.M. generated crucial reagents.

M.S.D planned experiments and analyzed data.

O.Z. designed the study, wrote the study protocol, processed PBMC samples, performed experiments, analyzed the data, and supervised the project.

B.J.L. planned the experiments, analyzed the data, prepared the manuscript, and supervised the project.

## COMPETING INTERESTS

M.S.D. is a consultant for Inbios, Vir Biotechnology, Senda Biosciences, and Carnival Corporation, and is on the Scientific Advisory Boards of Moderna and Immunome. The Diamond laboratory has received unrelated funding support in sponsored research agreements from Moderna, Vir Biotechnology, Immunome, and Emergent BioSolutions.

O.Z. and family own Moderna stock. The Ellebedy laboratory received unrelated funding support from Emergent BioSolutions and AbbVie.

A.H.E. is a consultant for Mubadala Investment Company and the founder of ImmuneBio Consulting. J.S.T. is a consultant for Gerson Lehrman Group.

J.S.T. and A.H.E. are recipients of a licensing agreement with Abbvie that is unrelated to this manuscript.

## Notes

### Author Declarations

The Institutional Review Board of Washington University School of Medicine gave ethical approval for this work (Approval #202104138 and 202012081).

## REFERENCES

1. L. R. Baden, H. M. E. Sahly, B. Essink, K. Kotloff, S. Frey, R. Novak, D. Diemert, S. A. Spector, N. Rouphael, C. B. Creech, J. McGettigan, S. Khetan, N. Segall, J. Solis, A. Brosz, C. Fierro, H. Schwartz, K. Neuzil, L. Corey, P. Gilbert, H. Janes, D. Follmann, M. Marovich, J. Mascola, L. Polakowski, J. Ledgerwood, B. S. Graham, H. Bennett, R. Pajon, C. Knightly, B. Leav, W. Deng, H. Zhou, S. Han, M. Ivarsson, J. Miller, T. Zaks, C. S. Group, Efficacy and Safety of the mRNA-1273 SARS-CoV-2 Vaccine. New Engl J Med. 384, 403–416 (2020).

2. F. P. Polack, S. J. Thomas, N. Kitchin, J. Absalon, A. Gurtman, S. Lockhart, J. L. Perez, G. P. Marc, E. D. Moreira, C. Zerbini, R. Bailey, K. A. Swanson, S. Roychoudhury, K. Koury, P. Li, W. V. Kalina, D. Cooper, R. W. Frenck, L. L. Hammitt, Ö. Türeci, H. Nell, A. Schaefer, S. Ünal, D. B. Tresnan, S. Mather, P. R. Dormitzer, U. Şahin, K. U. Jansen, W. C. Gruber, C. C. T. Group, Safety and Efficacy of the BNT162b2 mRNA Covid-19 Vaccine. New Engl J Med. 383, 2603–2615 (2020).

3. L. Tang, D. R. Hijano, A. H. Gaur, T. L. Geiger, E. J. Neufeld, J. M. Hoffman, R. T. Hayden, Asymptomatic and Symptomatic SARS-CoV-2 Infections After BNT162b2 Vaccination in a Routinely Screened Workforce. Jama. 325 (2021), doi:10.1001/jama.2021.6564.

4. Y. Angel, A. Spitzer, O. Henig, E. Saiag, E. Sprecher, H. Padova, R. Ben-Ami, Association Between Vaccination With BNT162b2 and Incidence of Symptomatic and Asymptomatic SARS-CoV-2 Infections Among Health Care Workers. Jama. 325 (2021), doi:10.1001/jama.2021.7152.

5. E. J. Haas, F. J. Angulo, J. M. McLaughlin, E. Anis, S. R. Singer, F. Khan, N. Brooks, M. Smaja, G. Mircus, K. Pan, J. Southern, D. L. Swerdlow, L. Jodar, Y. Levy, S. Alroy-Preis, Impact and effectiveness of mRNA BNT162b2 vaccine against SARS-CoV-2 infections and COVID-19 cases, hospitalisations, and deaths following a nationwide vaccination campaign in Israel: an observational study using national surveillance data. Lancet. 397, 1819–1829 (2021).

6. G. Regev-Yochay, S. Amit, M. Bergwerk, M. Lipsitch, E. Leshem, R. Kahn, Y. Lustig, C. Cohen, R. Doolman, A. Ziv, I. Novikov, C. Rubin, I. Gimpelevich, A. Huppert, G. Rahav, A. Afek, Y. Kreiss, Decreased infectivity following BNT162b2 vaccination: A prospective cohort study in Israel. Lancet Regional Heal - Europe. 7, 100150 (2021).

7. J. Sadoff, G. Gray, A. Vandebosch, V. Cárdenas, G. Shukarev, B. Grinsztejn, P. A. Goepfert, C. Truyers, H. Fennema, B. Spiessens, K. Offergeld, G. Scheper, K. L. Taylor, M. L. Robb, J. Treanor, D. H. Barouch, J. Stoddard, M. F. Ryser, M. A. Marovich, K. M. Neuzil, L. Corey, N. Cauwenberghs, T. Tanner, K. Hardt, J. Ruiz-Guiñazú, M. L. Gars, H. Schuitemaker, J. V. Hoof, F. Struyf, M. Douoguih, E. S. Group, Safety and Efficacy of Single-Dose Ad26.COV2.S Vaccine against Covid-19. New Engl J Medicine. 384, NEJMoa2101544 (2021).

8. W. Kim, J. Q. Zhou, S. C. Horvath, A. J. Schmitz, A. J. Sturtz, T. Lei, Z. Liu, E. Kalaidina, M. Thapa, W. B. Alsoussi, A. Haile, M. K. Klebert, T. Suessen, L. Parra-Rodriguez, P. A. Mudd, S. P. J. Whelan, W. D. Middleton, S. A. Teefey, I. Pusic, J. A. O’Halloran, R. M. Presti, J. S. Turner, A. H. Ellebedy, Germinal centre-driven maturation of B cell response to mRNA vaccination. Nature. 604, 141–145 (2022).

9. E. Cameroni, J. E. Bowen, L. E. Rosen, C. Saliba, S. K. Zepeda, K. Culap, D. Pinto, L. A. VanBlargan, A. D. Marco, J. di Iulio, F. Zatta, H. Kaiser, J. Noack, N. Farhat, N. Czudnochowski, C. Havenar-Daughton, K. R. Sprouse, J. R. Dillen, A. E. Powell, A. Chen, C. Maher, L. Yin, D. Sun, L. Soriaga, J. Bassi, C. Silacci-Fregni, C. Gustafsson, N. M. Franko, J. Logue, N. T. Iqbal, I. Mazzitelli, J. Geffner, R. Grifantini, H. Chu, A. Gori, A. Riva, O. Giannini, A. Ceschi, P. Ferrari, P. E. Cippà, A. Franzetti-Pellanda, C. Garzoni, P. J. Halfmann, Y. Kawaoka, C. Hebner, L. A. Purcell, L. Piccoli, M. S. Pizzuto, A. C. Walls, M. S. Diamond, A. Telenti, H. W. Virgin, A. Lanzavecchia, G. Snell, D. Veesler, D. Corti, Broadly neutralizing antibodies overcome SARS-CoV-2 Omicron antigenic shift. Nature. 602, 664–670 (2022).

10. K. Wang, Z. Jia, L. Bao, L. Wang, L. Cao, H. Chi, Y. Hu, Q. Li, Y. Zhou, Y. Jiang, Q. Zhu, Y. Deng, P. Liu, N. Wang, L. Wang, M. Liu, Y. Li, B. Zhu, K. Fan, W. Fu, P. Yang, X. Pei, Z. Cui, L. Qin, P. Ge, J. Wu, S. Liu, Y. Chen, W. Huang, Q. Wang, C.-F. Qin, Y. Wang, C. Qin, X. Wang, Memory B cell repertoire from triple vaccinees against diverse SARS-CoV-2 variants. Nature. 603, 919–925 (2022).

11. W. F. Garcia-Beltran, K. J. St. Denis, A. Hoelzemer, E. C. Lam, A. D. Nitido, M. L. Sheehan, C. Berrios, O. Ofoman, C. C. Chang, B. M. Hauser, J. Feldman, A. L. Roederer, D. J. Gregory, M. C. Poznansky, A. G. Schmidt, A. J. Iafrate, V. Naranbhai, A. B. Balazs, mRNA-based COVID-19 vaccine boosters induce neutralizing immunity against SARS-CoV-2 Omicron variant. Cell. 185, 457-466.e4 (2022).

12. A. Muik, A.-K. Wallisch, B. Sänger, K. A. Swanson, J. Mühl, W. Chen, H. Cai, D. Maurus, R. Sarkar, Ö. Türeci, P. R. Dormitzer, U. Şahin, Neutralization of SARS-CoV-2 lineage B.1.1.7 pseudovirus by BNT162b2 vaccine–elicited human sera. Science. 371, 1152–1153 (2021).

13. I. Meyts, G. Bucciol, I. Quinti, B. Neven, A. Fischer, E. Seoane, E. Lopez-Granados, C. Gianelli, A. Robles-Marhuenda, P.-Y. Jeandel, C. Paillard, V. G. Sankaran, Y. Y. Demirdag, V. Lougaris, A. Aiuti, A. Plebani, C. Milito, V. ASH. Dalm, K. Guevara-Hoyer, S. Sánchez-Ramón, L. Bezrodnik, F. Barzaghi, L. I. Gonzalez-Granado, G. R. Hayman, G. Uzel, L. O. Mendonça, C. Agostini, G. Spadaro, R. Badolato, A. Soresina, F. Vermeulen, C. Bosteels, B. N. Lambrecht, M. Keller, P. J. Mustillo, R. S. Abraham, S. Gupta, A. Ozen, E. Karakoc-Aydiner, S. Baris, A. F. Freeman, M. Yamazaki-Nakashimada, S. Scheffler-Mendoza, S. Espinosa-Padilla, A. R. Gennery, S. Jolles, Y. Espinosa, M. C. Poli, C. Fieschi, F. Hauck, C. Cunningham-Rundles, N. Mahlaoui, I. C. of I. E. of Immunity, K. Warnatz, K. E. Sullivan, S. G. Tangye, Coronavirus disease 2019 in patients with inborn errors of immunity: An international study. J Allergy Clin Immunol. 147, 520–531 (2021).

14. A. M. Shields, A. Anantharachagan, G. Arumugakani, K. Baker, S. Bahal, H. Baxendale, W. Bermingham, M. Bhole, E. Boules, P. Bright, C. Chopra, L. Cliffe, B. Cleave, J. Dempster, L. Devlin, F. Dhalla, L. Diwakar, E. Drewe, C. Duncan, M. Dziadzio, S. Elcombe, S. Elkhalifa, A. Gennery, H. Ghanta, S. Goddard, S. Grigoriadou, S. Hackett, G. Hayman, R. Herriot, A. Herwadkar, A. Huissoon, R. Jain, S. Jolles, S. Johnston, S. Khan, J. Laffan, P. Lane, L. Leeman, D. M. Lowe, S. Mahabir, D. J. M. Lochlainn, E. McDermott, S. Misbah, F. Moghaddas, H. Morsi, S. Murng, S. Noorani, R. O’Brien, S. Patel, A. Price, T. Rahman, S. Seneviratne, A. Shrimpton, C. Stroud, M. Thomas, K. Townsend, P. Vaitla, N. Verma, A. Williams, S. O. Burns, S. Savic, A. G. Richter, Outcomes following SARS-CoV-2 infection in patients with primary and secondary immunodeficiency in the United Kingdom. Clin Exp Immunol, uxac008 (2022).

15. D. Arroyo-Sánchez, O. Cabrera-Marante, R. Laguna-Goya, P. Almendro-Vázquez, O. Carretero, F. J. Gil-Etayo, P. Suàrez-Fernández, P. Pérez-Romero, E. R. de A. Serrano, L. M. Allende, D. Pleguezuelo, E. Paz-Artal, Immunogenicity of Anti-SARS-CoV-2 Vaccines in Common Variable Immunodeficiency. J Clin Immunol. 42, 240–252 (2022).

16. S. A. Apostolidis, M. Kakara, M. M. Painter, R. R. Goel, D. Mathew, K. Lenzi, A. Rezk, K. R. Patterson, D. A. Espinoza, J. C. Kadri, D. M. Markowitz, C. E. Markowitz, I. Mexhitaj, D. Jacobs, A. Babb, M. R. Betts, E. T. L. Prak, D. Weiskopf, A. Grifoni, K. A. Lundgreen, S. Gouma, A. Sette, P. Bates, S. E. Hensley, A. R. Greenplate, E. J. Wherry, R. Li, A. Bar-Or, Cellular and humoral immune responses following SARS-CoV-2 mRNA vaccination in patients with multiple sclerosis on anti-CD20 therapy. Nat Med. 27, 1990–2001 (2021).

17. E. Azzolini, C. Pozzi, L. Germagnoli, B. Oresta, N. Carriglio, M. Calleri, C. Selmi, M. D. Santis, S. Finazzi, C. Carlo-Stella, A. Bertuzzi, F. Motta, A. Ceribelli, A. Mantovani, F. Bonelli, M. Rescigno, mRNA COVID-19 vaccine booster fosters B- and T-cell responses in immunocompromised patients. Life Sci Alliance. 5, e202201381 (2022).

18. N. A. Kennedy, S. Lin, J. R. Goodhand, N. Chanchlani, B. Hamilton, C. Bewshea, R. Nice, D. Chee, J. F. Cummings, A. Fraser, P. M. Irving, N. Kamperidis, K. B. Kok, C. A. Lamb, J. Macdonald, S. Mehta, R. C. Pollok, T. Raine, P. J. Smith, A. M. Verma, S. Jochum, T. J. McDonald, S. Sebastian, C. W. Lees, N. Powell, T. Ahmad C. to the C. I. study, Gut, in press, doi:10.1136/gutjnl-2021-324789.

19. S. K. Mahil, K. Bechman, A. Raharja, C. Domingo-Vila, D. Baudry, M. A. Brown, A. P. Cope, T. Dasandi, C. Graham, T. Lechmere, M. H. Malim, F. Meynell, E. Pollock, J. Seow, K. Sychowska, J. N. Barker, S. Norton, J. B. Galloway, K. J. Doores, T. I. M. Tree, C. H. Smith, The effect of methotrexate and targeted immunosuppression on humoral and cellular immune responses to the COVID-19 vaccine BNT162b2: a cohort study. Lancet Rheumatology. 3, e627–e637 (2021).

20. T. Bachelet, J.-P. Bourdenx, C. Martinez, S. Mucha, P. Martin-Dupont, V. Perier, A. Pommereau, Humoral response after SARS-CoV-2 mRNA vaccines in dialysis patients: Integrating anti-SARS-CoV-2 Spike-Protein-RBD antibody monitoring to manage dialysis centers in pandemic times. Plos One. 16, e0257646 (2021).

21. C. Danthu, S. Hantz, A. Dahlem, M. Duval, B. Ba, M. Guibbert, Z. E. Ouafi, S. Ponsard, I. Berrahal, J. M. Achard, F. Bocquentin, V. Allot, J. P. Rerolle, S. Alain, F. Toure, J Am Soc Nephrol, in press, doi:10.1681/asn.2021040490.

22. P. Deepak, W. Kim, M. A. Paley, M. Yang, A. B. Carvidi, E. G. Demissie, A. A. El-Qunni, A. Haile, K. Huang, B. Kinnett, M. J. Liebeskind, Z. Liu, L. E. McMorrow, D. Paez, N. Pawar, D. C. Perantie, R. E. Schriefer, S. E. Sides, M. Thapa, M. Gergely, S. Abushamma, S. Akuse, M. Klebert, L. Mitchell, D. Nix, J. Graf, K. E. Taylor, S. Chahin, M. A. Ciorba, P. Katz, M. Matloubian, J. A. O’Halloran, R. M. Presti, G. F. Wu, S. P. J. Whelan, W. J. Buchser, L. S. Gensler, M. C. Nakamura, A. H. Ellebedy, A. H. J. Kim, Effect of Immunosuppression on the Immunogenicity of mRNA Vaccines to SARS-CoV-2. Ann Intern Med. 174, M21–1757 (2021).

23. L. P. M. van Leeuwen, C. H. GeurtsvanKessel, P. M. Ellerbroek, G. J. de Bree, J. Potjewijd, A. Rutgers, H. Jolink, F. van de Veerdonk, E. C. M. van Gorp, F. de Wilt, S. Bogers, L. Gommers, D. Geers, A. H. W. Bruns, H. L. Leavis, J. W. van Haga, B. A. Lemkes, A. van der Veen, S. F. J. de Kruijf-Bazen, P. van Paassen, K. de Leeuw, A. A. J. M. van de Ven, P. H. Verbeek-Menken, A. van Wengen, S. M. Arend, A. J. Ruten-Budde, M. W. van der Ent, P. M. van Hagen, R. W. Sanders, M. Grobben, K. van der Straten, J. A. Burger, M. Poniman, S. Nierkens, M. J. van Gils, R. D. de Vries, V. A. S. H. Dalm, Immunogenicity of the mRNA-1273 COVID-19 vaccine in adult patients with inborn errors of immunity. J Allergy Clin Immunol (2022), doi:10.1016/j.jaci.2022.04.002.

24. O. Zimmerman, A. M. A. Doss, P. Kaplonek, C.-Y. Liang, L. A. VanBlargan, R. E. Chen, J. M. Monroy, H. J. Wedner, A. Kulczycki, T. L. Mantia, C. C. O’Shaughnessy, H. G. Davis-Adams, H. L. Bertera, L. J. Adams, S. Raju, F. R. Zhao, C. J. Rigell, T. B. Dy, A. L. Kau, Z. Ren, J. S. Turner, J. A. O’Halloran, R. M. Presti, D. H. Fremont, P. L. Kendall, A. H. Ellebedy, G. Alter, M. S. Diamond, mRNA vaccine boosting enhances antibody responses against SARS-CoV-2 Omicron variant in individuals with antibody deficiency syndromes. Cell Reports Medicine, 100653 (2022).

25. A. Durandy, S. Kracker, A. Fischer, Primary antibody deficiencies. Nat Rev Immunol. 13, 519–533 (2013).

26. H. Abolhassani, L. Hammarström, C. Cunningham-Rundles, Current genetic landscape in common variable immune deficiency. Blood. 135, 656–667 (2020).

27. G. de Valles-Ibáñez, A. Esteve-Solé, M. Piquer, E. A. González-Navarro, J. Hernandez-Rodriguez, H. Laayouni, E. González-Roca, A. M. Plaza-Martin, Á. Deyà-Martínez, A. Martín-Nalda, M. Martínez-Gallo, M. García-Prat, L. del Pino-Molina, I. Cuscó, M. Codina-Solà, L. Batlle-Masó, M. Solís-Moruno, T. Marquès-Bonet, E. Bosch, E. López-Granados, J. I. Aróstegui, P. Soler-Palacín, R. Colobran, J. Yagüe, L. Alsina, M. Juan, F. Casals, Evaluating the Genetics of Common Variable Immunodeficiency: Monogenetic Model and Beyond. Front Immunol. 9, 636 (2018).

28. D. J. A. Bogaert, M. Dullaers, B. N. Lambrecht, K. Y. Vermaelen, E. D. Baere, F. Haerynck, Genes associated with common variable immunodeficiency: one diagnosis to rule them all? J Med Genet. 53, 575 (2016).

29. P. Maffucci, C. A. Filion, B. Boisson, Y. Itan, L. Shang, J.-L. Casanova, C. Cunningham-Rundles, Genetic Diagnosis Using Whole Exome Sequencing in Common Variable Immunodeficiency. Front Immunol. 7, 220 (2016).

30. P. Tuijnenburg, H. L. Allen, S. O. Burns, D. Greene, M. H. Jansen, E. Staples, J. Stephens, K. J. Carss, D. Biasci, H. Baxendale, M. Thomas, A. Chandra, S. Kiani-Alikhan, H. J. Longhurst, S. L. Seneviratne, E. Oksenhendler, I. Simeoni, G. J. de Bree, A. T. J. Tool, E. M. M. van Leeuwen, E. H. T. M. Ebberink, A. B. Meijer, S. Tuna, D. Whitehorn, M. Brown, E. Turro, A. J. Thrasher, K. G. C. Smith, J. E. Thaventhiran, T. W. Kuijpers, Z. Adhya, H. Alachkar, A. Anantharachagan, R. Antrobus, G. Arumugakani, C. Bacchelli, H. Baxendale, C. Bethune, S. Bibi, B. Boardman, C. Booth, M. Browning, M. Brownlie, S. Burns, A. Chandra, H. Clifford, N. Cooper, S. Davies, J. Dempster, L. Devlin, R. Doffinger, E. Drewe, D. Edgar, W. Egner, T. El-Shanawany, B. Gaspar, R. Ghurye, K. Gilmour, S. Goddard, P. Gordins, S. Grigoriadou, S. Hackett, R. Hague, L. Harper, G. Hayman, A. Herwadkar, S. Hughes, A. Huissoon, S. Jolles, J. Jones, P. Kelleher, N. Klein, T. Kuijpers, D. Kumararatne, J. Laffan, H. L. Allen, S. Lear, H. Longhurst, L. Lorenzo, J. Maimaris, A. Manson, E. McDermott, H. Millar, A. Mistry, V. Morrisson, S. Murng, I. Nasir, S. Nejentsev, S. Noorani, E. Oksenhendler, M. Ponsford, W. Qasim, E. Quinn, I. Quinti, A. Richter, C. Samarghitean, R. Sargur, S. Savic, S. Seneviratne, C. Sewall, F. Shackley, I. Simeoni, K. G. C. Smith, E. Staples, H. Stauss, C. Steele, J. Thaventhiran, M. Thomas, A. Thrasher, S. Welch, L. Willcocks, S. Workman, A. Worth, N. Yeatman, P. Yong, S. Ashford, J. Bradley, D. Fletcher, T. Hammerton, R. James, N. Kingston, W. Ouwehand, C. Penkett, F. L. Raymond, K. Stirrups, M. Veltman, T. Young, S. Ashford, M. Brown, N. Clements-Brod, J. Davis, E. Dewhurst, M. Erwood, A. Frary, R. Linger, J. Martin, S. Papadia, K. Rehnstrom, W. Astle, A. Attwood, M. Bleda, K. Carss, L. Daugherty, S. Deevi, S. Graf, D. Greene, C. Halmagyi, M. Haimel, F. Hu, R. James, H. L. Allen, V. Matser, S. Meacham, K. Megy, C. Penkett, O. Shamardina, K. Stirrups, C. Titterton, S. Tuna, E. Turro, P. Yu, J. von Ziegenweldt, A. Furnell, R. Mapeta, I. Simeoni, S. Staines, J. Stephens, K. Stirrups, D. Whitehorn, P. Rayner-Matthews, C. Watt, Loss-of-function nuclear factor κB subunit 1 (NFKB1) variants are the most common monogenic cause of common variable immunodeficiency in Europeans. J Allergy Clin Immunol. 142, 1285–1296 (2018).

31. L. J. Maarschalk-Ellerbroek, I. M. Hoepelman, P. M. Ellerbroek, Immunoglobulin treatment in primary antibody deficiency. Int J Antimicrob Ag. 37, 396–404 (2011).

32. J. Prevot, S. Jolles, Global immunoglobulin supply: steaming towards the iceberg? Curr Opin Allergy Cl. 20, 557–564 (2020).

33. A. L. Miller, N. L. Rider, R. B. Pyles, B. Judy, X. Xie, P.-Y. Shi, T. G. Ksiazek, The arrival of SARS-CoV-2–neutralizing antibodies in a currently available commercial immunoglobulin. J Allergy Clin Immunol. 149, 1958–1959 (2022).

34. W. B. Alsoussi, J. S. Turner, J. B. Case, H. Zhao, A. J. Schmitz, J. Q. Zhou, R. E. Chen, T. Lei, A. A. Rizk, K. M. McIntire, E. S. Winkler, J. M. Fox, N. M. Kafai, L. B. Thackray, A. O. Hassan, F. Amanat, F. Krammer, C. T. Watson, S. H. Kleinstein, D. H. Fremont, M. S. Diamond, A. H. Ellebedy, A Potently Neutralizing Antibody Protects Mice against SARS-CoV-2 Infection. J Immunol. 205, ji2000583 (2020).

35. A. O. Hassan, J. B. Case, E. S. Winkler, L. B. Thackray, N. M. Kafai, A. L. Bailey, B. T. McCune, J. M. Fox, R. E. Chen, W. B. Alsoussi, J. S. Turner, A. J. Schmitz, T. Lei, S. Shrihari, S. P. Keeler, D. H. Fremont, S. Greco, P. B. McCray, S. Perlman, M. J. Holtzman, A. H. Ellebedy, M. S. Diamond, A SARS-CoV-2 Infection Model in Mice Demonstrates Protection by Neutralizing Antibodies. Cell. 182, 744-753.e4 (2020).

36. G. E. Weiss, P. D. Crompton, S. Li, L. A. Walsh, S. Moir, B. Traore, K. Kayentao, A. Ongoiba, O. K. Doumbo, S. K. Pierce, Atypical Memory B Cells Are Greatly Expanded in Individuals Living in a Malaria-Endemic Area. J Immunol. 183, 2176–2182 (2009).

37. H. J. Sutton, R. Aye, A. H. Idris, R. Vistein, E. Nduati, O. Kai, J. Mwacharo, X. Li, X. Gao, T. D. Andrews, M. Koutsakos, T. H. O. Nguyen, M. Nekrasov, P. Milburn, A. Eltahla, A. A. Berry, N. Kc, S. Chakravarty, B. K. L. Sim, A. K. Wheatley, S. J. Kent, S. L. Hoffman, K. E. Lyke, P. Bejon, F. Luciani, K. Kedzierska, R. A. Seder, F. M. Ndungu, I. A. Cockburn, Atypical B cells are part of an alternative lineage of B cells that participates in responses to vaccination and infection in humans. Cell Reports. 34, 108684 (2021).

38. M.-L. Golinski, M. Demeules, C. Derambure, G. Riou, M. Maho-Vaillant, O. Boyer, P. Joly, S. Calbo, CD11c+ B Cells Are Mainly Memory Cells, Precursors of Antibody Secreting Cells in Healthy Donors. Front Immunol. 11, 32 (2020).

39. H. Rincon-Arevalo, A. Wiedemann, A.-L. Stefanski, M. Lettau, F. Szelinski, S. Fuchs, A. P. Frei, M. Steinberg, T. Kam-Thong, K. Hatje, B. Keller, K. Warnatz, A. Radbruch, A. C. Lino, E. Schrezenmeier, T. Dörner, Deep Phenotyping of CD11c+ B Cells in Systemic Autoimmunity and Controls. Front Immunol. 12, 635615 (2021).

40. S. Shalapour, J. Font-Burgada, G. D. Caro, Z. Zhong, E. Sanchez-Lopez, D. Dhar, G. Willimsky, M. Ammirante, A. Strasner, D. E. Hansel, C. Jamieson, C. J. Kane, T. Klatte, P. Birner, L. Kenner, M. Karin, Immunosuppressive plasma cells impede T-cell-dependent immunogenic chemotherapy. Nature. 521, 94–98 (2019).

41. P. A. Mudd, A. A. Minervina, M. V. Pogorelyy, J. S. Turner, W. Kim, E. Kalaidina, J. Petersen, A. J. Schmitz, T. Lei, A. Haile, A. M. Kirk, R. C. Mettelman, J. C. Crawford, T. H. O. Nguyen, L. C. Rowntree, E. Rosati, K. A. Richards, A. J. Sant, M. K. Klebert, T. Suessen, W. D. Middleton, S. S. Team, J. H. Estepp, S. Schultz-Cherry, M. A. McGargill, A. Gaur, J. Hoffman, M. Mori, L. Tang, E. Tuomanen, R. Webby, R. T. Hayden, H. Hakim, D. R. Hijano, K. J. Allison, E. K. Allen, R. Bajracharya, W. Awad, L.-A. V. de Velde, B. L. Clark, T. L. Wilson, A. Souquette, A. Castellaw, R. H. Dallas, A. Gowen, T. P. Fabrizio, C.-Y. Lin, D. C. Brice, S. Cherry, E. K. Roubidoux, V. Cortez, P. Freiden, N. Wohlgemuth, K. Whitt, J. Wolf, S. A. Teefey, J. A. O’Halloran, R. M. Presti, K. Kedzierska, J. Rossjohn, P. G. Thomas, A. H. Ellebedy, SARS-CoV-2 mRNA vaccination elicits a robust and persistent T follicular helper cell response in humans. Cell. 185, 603-613.e15 (2022).

42. L. Loyal, J. Braun, L. Henze, B. Kruse, M. Dingeldey, U. Reimer, F. Kern, T. Schwarz, M. Mangold, C. Unger, F. Dörfler, S. Kadler, J. Rosowski, K. Gürcan, Z. Uyar-Aydin, M. Frentsch, F. Kurth, K. Schnatbaum, M. Eckey, S. Hippenstiel, A. Hocke, M. A. Müller, B. Sawitzki, S. Miltenyi, F. Paul, M. A. Mall, H. Wenschuh, S. Voigt, C. Drosten, R. Lauster, N. Lachman, L.-E. Sander, V. M. Corman, J. Röhmel, L. Meyer-Arndt, A. Thiel, C. Giesecke-Thiel, Cross-reactive CD4+ T cells enhance SARS-CoV-2 immune responses upon infection and vaccination. Science. 374, eabh1823 (2021).

43. A. G. Dykema, B. Zhang, B. A. Woldemeskel, C. C. Garliss, L. S. Cheung, D. Choudhury, J. Zhang, L. Aparicio, S. Bom, R. Rashid, J. X. Caushi, E. H.-C. Hsiue, K. Cascino, E. A. Thompson, A. K. Kwaa, D. Singh, S. Thapa, A. A. Ordonez, A. Pekosz, F. R. D’Alessio, J. D. Powell, S. Yegnasubramanian, S. Zhou, D. M. Pardoll, H. Ji, A. L. Cox, J. N. Blankson, K. N. Smith, Functional characterization of CD4+ T-cell receptors cross-reactive for SARS-CoV-2 and endemic coronaviruses. J Clin Invest. 131 (2021), doi:10.1172/jci146922.

44. B. J. Laidlaw, A. H. Ellebedy, The germinal centre B cell response to SARS-CoV-2. Nat Rev Immunol. 22, 7–18 (2022).

45. R. R. Goel, S. A. Apostolidis, M. M. Painter, D. Mathew, A. Pattekar, O. Kuthuru, S. Gouma, P. Hicks, W. Meng, A. M. Rosenfeld, S. Dysinger, K. A. Lundgreen, L. Kuri-Cervantes, S. Adamski, A. Hicks, S. Korte, D. A. Oldridge, A. E. Baxter, J. R. Giles, M. E. Weirick, C. M. McAllister, J. Dougherty, S. Long, K. D’Andrea, J. T. Hamilton, M. R. Betts, E. T. L. Prak, P. Bates, S. E. Hensley, A. R. Greenplate, E. J. Wherry, Distinct antibody and memory B cell responses in SARS-CoV-2 naïve and recovered individuals following mRNA vaccination. Sci Immunol. 6, eabi6950 (2021).

46. F. Muecksch, Z. Wang, A. Cho, C. Gaebler, T. B. Tanfous, J. DaSilva, E. Bednarski, V. Ramos, S. Zong, B. Johnson, R. Raspe, D. Schaefer-Babajew, I. Shimeliovich, M. Daga, K.-H. Yao, F. Schmidt, K. G. Millard, M. Turroja, M. Jankovic, T. Y. Oliveira, A. Gazumyan, M. Caskey, T. Hatziioannou, P. D. Bieniasz, M. C. Nussenzweig, Increased memory B cell potency and breadth after a SARS-CoV-2 mRNA boost. Nature, 1–7 (2022).

47. A. F. Salinas, E. P. Mortari, S. Terreri, C. Quintarelli, F. Pulvirenti, S. D. Cecca, M. Guercio, C. Milito, L. Bonanni, S. Auria, L. Romaggioli, G. Cusano, C. Albano, S. Zaffina, C. F. Perno, G. Spadaro, F. Locatelli, R. Carsetti, I. Quinti, SARS-CoV-2 Vaccine Induced Atypical Immune Responses in Antibody Defects: Everybody Does their Best. J Clin Immunol. 41, 1709–1722 (2021).

48. F. Pulvirenti, A. F. Salinas, C. Milito, S. Terreri, E. P. Mortari, C. Quintarelli, S. D. Cecca, G. Lagnese, A. Punziano, M. Guercio, L. Bonanni, S. Auria, F. Villani, C. Albano, F. Locatelli, G. Spadaro, R. Carsetti, I. Quinti, B Cell Response Induced by SARS-CoV-2 Infection Is Boosted by the BNT162b2 Vaccine in Primary Antibody Deficiencies. Cells. 10, 2915 (2021).

49. J. B. Case, S. Mackin, J. Errico, Z. Chong, E. A. Madden, B. Guarino, M. A. Schmid, K. Rosenthal, K. Ren, A. Jung, L. Droit, S. A. Handley, P. J. Halfmann, Y. Kawaoka, J. E. Crowe, D. H. Fremont, H. W. Virgin, Y.-M. Loo, M. T. Esser, L. A. Purcell, D. Corti, M. S. Diamond, Biorxiv, in press, doi:10.1101/2022.03.17.484787.

50. J. S. Turner, J. A. O’Halloran, E. Kalaidina, W. Kim, A. J. Schmitz, J. Q. Zhou, T. Lei, M. Thapa, R. E. Chen, J. B. Case, F. Amanat, A. M. Rauseo, A. Haile, X. Xie, M. K. Klebert, T. Suessen, W. D. Middleton, P.-Y. Shi, F. Krammer, S. A. Teefey, M. S. Diamond, R. M. Presti, A. H. Ellebedy, SARS-CoV-2 mRNA vaccines induce persistent human germinal centre responses. Nature. 596, 109–113 (2021).

