## Supplementary Information for "SARS-CoV-2 booster vaccination rescues attenuated IgG1 memory B cell response in primary antibody deficiency patients"

**This PDF includes:**

Supplementary methods

Supplementary references

Fig. S1. Baseline phenotype of PAD patients and healthy donor cohorts.
Fig S2. COVID-19-experienced PAD patients display elevated RBD-specific B cell response following primary vaccination series.
Fig. S3. COVID-19-naive PAD patients display elevated RBD-specific B cell response following booster vaccination.
Fig. S4. Isotype composition of SARS-CoV-2 RBD-specific memory B cell response following vaccination in PAD patients.
Fig. S5. RBD-specific memory B cells from PAD patients display reduced CD11c expression class.
Table S1. Characteristics of patient cohort
Table S2. Cellular phenotype of patient cohort
Table S3. PAD patient laboratory values
Table S4. PAD patient responses to other vaccine antigens at time of diagnosis

### SUPPLEMENTARY METHODS

**Preparation of peripheral blood mononuclear cells**. Patient blood was collected in sodium citrate cell preparation tubes (BD Biosciences). Tubes were centrifuged at room temperature for 30 minutes at 1650 x *g* without brake. The mononuclear cells and plasma layer were then transferred to a 50mL conical and centrifuged at room temperature for 10 minutes at 256 x *g*. Supernatant was removed without disturbing the cell pellet. Cells were resuspended in PBS and all cells of the same subject were combined into one 15-mL conical tube. Cells were centrifuged as above. Supernatant was removed without disturbing the cell pellet and cells were resuspended in 5mL of ACK lysing buffer (Thermo Fisher) for 5 minutes. Cells were washed with PBS, counted, washed again with PBS, resuspended in 10% dimethylsulfoxide in FBS at $10^6$ cells per mL and aliquoted into cryovials. Cryovials were transferred to a freezing container (Daigger Scientific, Mr. Frosty) and placed in a -80°C freezer overnight before being transferred to liquid nitrogen for storage.

**Antibodies for flow cytometry staining**. Staining for flow cytometry was performed using cryopreserved PBMC samples. Cyropreserved samples were thawed in a 37 °C water bath and added to tube containing RPMI with 10% FBS and 1% Penicillin Streptomycin solution (Fisher). Cells were then centrifuged at 300x *g* at room temperature, resuspended in 10% RPMI, and counted. Cells were then aliquoted to a 96 well plate and centrifuged at 810 x *g* for 2 minutes at 4 °C. The cells were then washed with PBS containing 2% FBS, centrifuged, and then incubated with the primary antibody mix for 30 minutes on ice. Samples that were stained with tetramer containing antibody cocktail were incubated for 45 minutes. Cells were then washed three times with 2% PBS and incubated with secondary antibody mix for an additional 30 minutes. Samples were then washed an additional three times with 2% PBS before being resuspended for flow cytometry analysis. Flow cytometry data were acquired on a Cytek Aurora and were analyzed with FlowJo software (TreeStar).

The following antibodies were used for flow cytometry staining: Brilliant Violet 711(BV711) anti-CD11c (301630), BV750 anti-CD19 (302262), allophycocyanin (APC)/Fire 810 anti-CD3 (344858), BV510 anti-IgD (348220), BV605 anti-IgM (314524), phycoerythrin (PE)/Dazzle 594 anti-CXCR5 (356928), APC anti-his (362605), PE/Fire 810 anti-CD27 (302859), APC/Fire 750 anti-CD20 (302358), Zombie NIR (423106), phycoerythrin-indotricarbocyanine (PE-Cy7) anti-CD71 (334112), BV650 BV570 anti-CD45RO (304226), FITC anti-CCR7 (353216), BV605 anti-

HLA-DR (307640), Alexa Fluor 700 anti-CD4 (344622), Alexa Fluor 594 anti-CD8 (301056), peridinin chlorophyll protein Cy5.5 (PerCpCy5.5) (304122), PE/Fire 810 anti-CD27 (302859), BV421 anti-ICOS (313524) (all from Biolegend); Biotin anti-IgG3 (OB9210-08), Alexa Fluor 555 anti-IgG2 (OB907032), PE anti-IgG1 (OB905409), FITC anti-IgA (CBL114FMI), Brilliant Blue 700 (BB700) anti-CD38 (BDB566445), PE-Cy7 anti-PD1 (BDB561272) (all from Fisher).

**His-tagged SARS-CoV-2 protein purification**. Genes encoding SARS-CoV-2 Wuhan-Hu-1 spike protein (residues 1-1213, GenBank: MN908947.3), the Wuhan-Hu-1 RBD (residues 319-514), and B.1.1.529 (BA.1) spike protein were cloned into a pCAGGS mammalian expression vector with a C-terminal hexahistidine tag. Both spike proteins were prefusion stabilized and expression optimized with six proline substitutions (F817P, A892P, A899P, A942P, K986P, V987P), with a disrupted S1/S2 furin cleavage site and a C-terminal foldon trimerization motif (YIPEAPRDGQAYVRKDGEWVLLSTFL) (*50*). Expi293F cells were transiently transfected, and proteins were recovered via cobalt-charged resin chromatography (G-Biosciences) as previously described (*34*, *35*) .

**HLA class II tetramers.** HLA class II tetramers representing the HLA-DPA1*01:03/HLA-DPB1*04:01-restricted SARS-CoV-2 spike (S) protein epitopes $S_{167-180}$ (TFEYVSQPFLMDLE) and $S_{816-830}$ (SFIEDLLFNKVTLAD) were constructed by cloning the relevant peptides into an AbVec vector system containing the appropriate HLA alpha and beta chains linked to the peptide of interest via a flexible linker at the N-terminus of the beta chain (*41–43*). This system includes corresponding leucine zipper motifs on the alpha and beta chains to promote HLA monomer stability. Proteins were expressed in 293F cells transfected with the appropriate AbVec vector. Purified HLA monomer was biotinylated with BioA enzyme in biotinylation buffer (0.2 M NaCl, 0.1 M Tris pH7.5, 5 mM $MgCl_2$, 5 mM ATP, 0.4 mM Biotin, 5 uM Leupeptin, 1 uM Pepstatin, and 0.2 mM PMSF). The monomer was then tetramerized and fluorochrome labeled by adding Streptavidin-R-Phycoerythrin (Agilent) to the $S_{167-180}$ monomer or Streptavidin-Allophycocyanin (Agilent) to the $S_{816-830}$ monomer (*51*).

**Focus reduction neutralization test.** Neutralizing antibody titers were performed with authentic SARS-CoV-2 strains and variants as previous described (*24*).

### SUPPLEMENTAL REFERENCES

51. C.-L. Hsieh, J. A. Goldsmith, J. M. Schaub, A. M. DiVenere, H.-C. Kuo, K. Javanmardi, K. C. Le, D. Wrapp, A. G. Lee, Y. Liu, C.-W. Chou, P. O. Byrne, C. K. Hjorth, N. V. Johnson, J. Ludes-Meyers, A. W. Nguyen, J. Park, N. Wang, D. Amengor, J. J. Lavinder, G. C. Ippolito, J. A. Maynard, I. J. Finkelstein, J. S. McLellan, Structure-based design of prefusion-stabilized SARS-CoV-2 spikes. *Science*. 369, eabd0826 (2020).

52. R. A. Willis, V. Ramachandiran, J. C. Shires, G. Bai, K. Jeter, D. L. Bell, L. Han, T. Kazarian, K. C. Ugwu, O. Laur, S. Contreras-Alcantara, D. L. Long, J. D. Altman, Production of Class II MHC Proteins in Lentiviral Vector-Transduced HEK-293T Cells for Tetramer Staining Reagents. *Curr Protoc*. 1, e36 (2021).

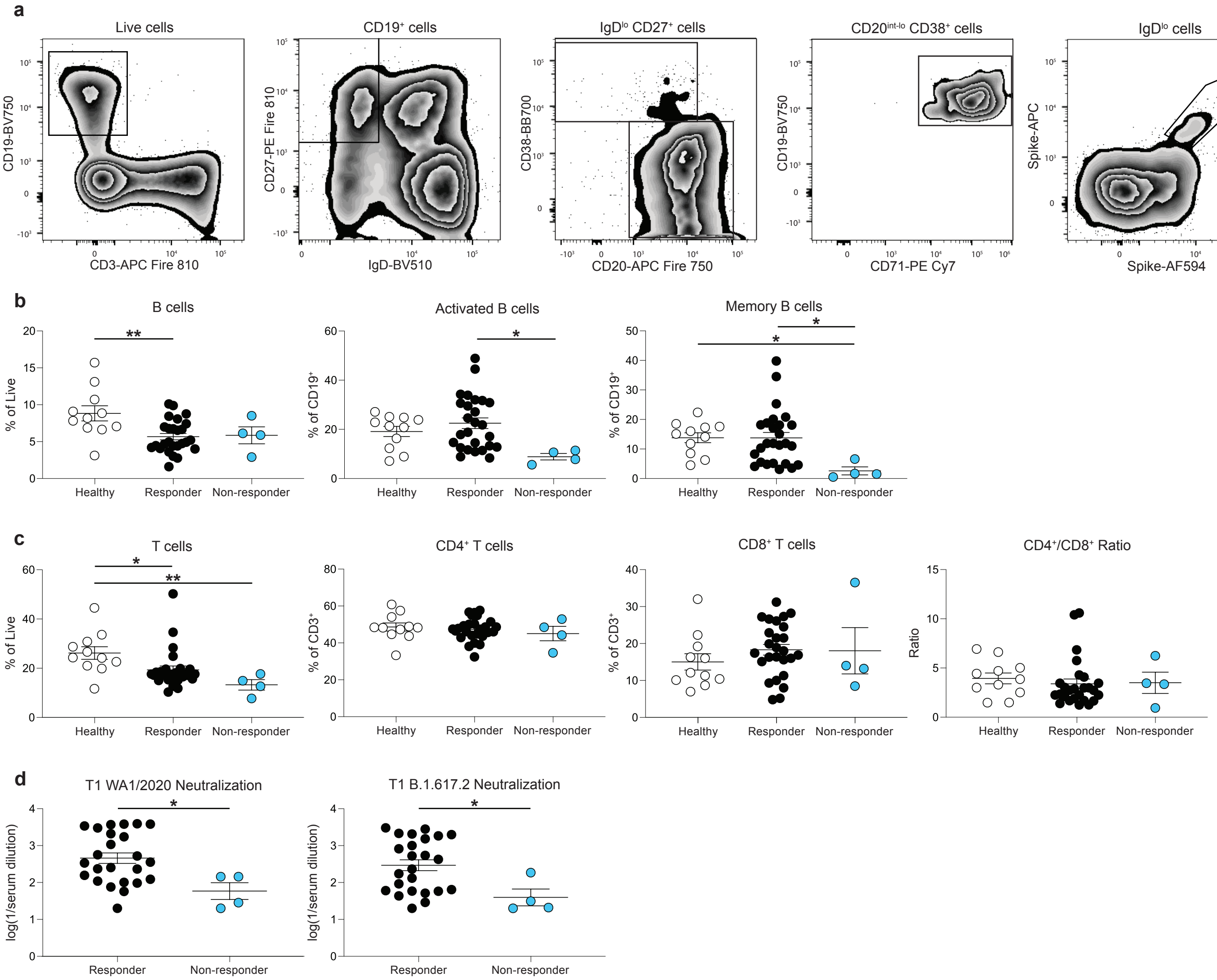
a
Live cells
CD19-BV750
CD3-APC Fire 810
CD19⁺ cells
CD27-PE Fire 810
IgD-BV510
IgDˡᵒ CD27⁺ cells
CD38-BB700
CD20-APC Fire 750
CD20ᶦⁿᵗ⁻ˡᵒ CD38⁺ cells
CD19-BV750
CD71-PE Cy7
IgDˡᵒ cells
Spike-APC
Spike-AF594
IgDˡᵒ cells
RBD-APC
RBD-AF594
b
B cells
% of Live
Activated B cells
% of CD19⁺
Memory B cells
% of CD19⁺
Healthy
Responder
Non-responder
c
T cells
% of Live
CD4⁺ T cells
% of CD3⁺
CD8⁺ T cells
% of CD3⁺
CD4⁺/CD8⁺ Ratio
Ratio
d
T1 WA1/2020 Neutralization
log(1/serum dilution)
T1 B.1.617.2 Neutralization
log(1/serum dilution)
Responder
Non-responder

**Figure S1**

**Figure S1**. **Baseline phenotype of PAD patients and healthy donor cohorts**. (**a**) Representative B cell flow cytometry gating scheme. (**b**) Percentage of live cells that are B ($CD19^+CD3^-$) cells at T1 in patient cohorts (left). Percentage of B cells that are activated ($IgD^{lo}$) cells in patient cohorts (middle) Percentage of B cells that are memory ($IgD^{lo}CD27^+$) cells in patient cohorts. (right). (**c**) Percentage of live cells that are T ($CD3^+CD19^-$) cells at T1 in patient cohorts. (left) Percentage of T cells that are $CD4^+$ ($CD4^+CD8^-$) cells in patient cohorts (middle left). Percentage of T cells that are $CD8^+$ ($CD8^+CD4^-$) in patient cohorts (middle right). Ratio of $CD4^+$ to $CD8^+$ T cells (right). (**d**) Serum neutralizing activity against WA1/2020 (left) and B.1.617.2 (right) at T1. Non-responder defined as PAD patients in which the $CD19^+$ $IgD^{lo}$ $Spike^+$ B cells response was <0.02% of total B cells at T1. Statistical analyses were performed using a one-way ANOVA with Fisher's least significant difference testing in b, c; or using an unpaired t-test in d. Error bars were calculated based on the standard error of the mean. (*, $p < 0.05$; **, $p < 0.01$).

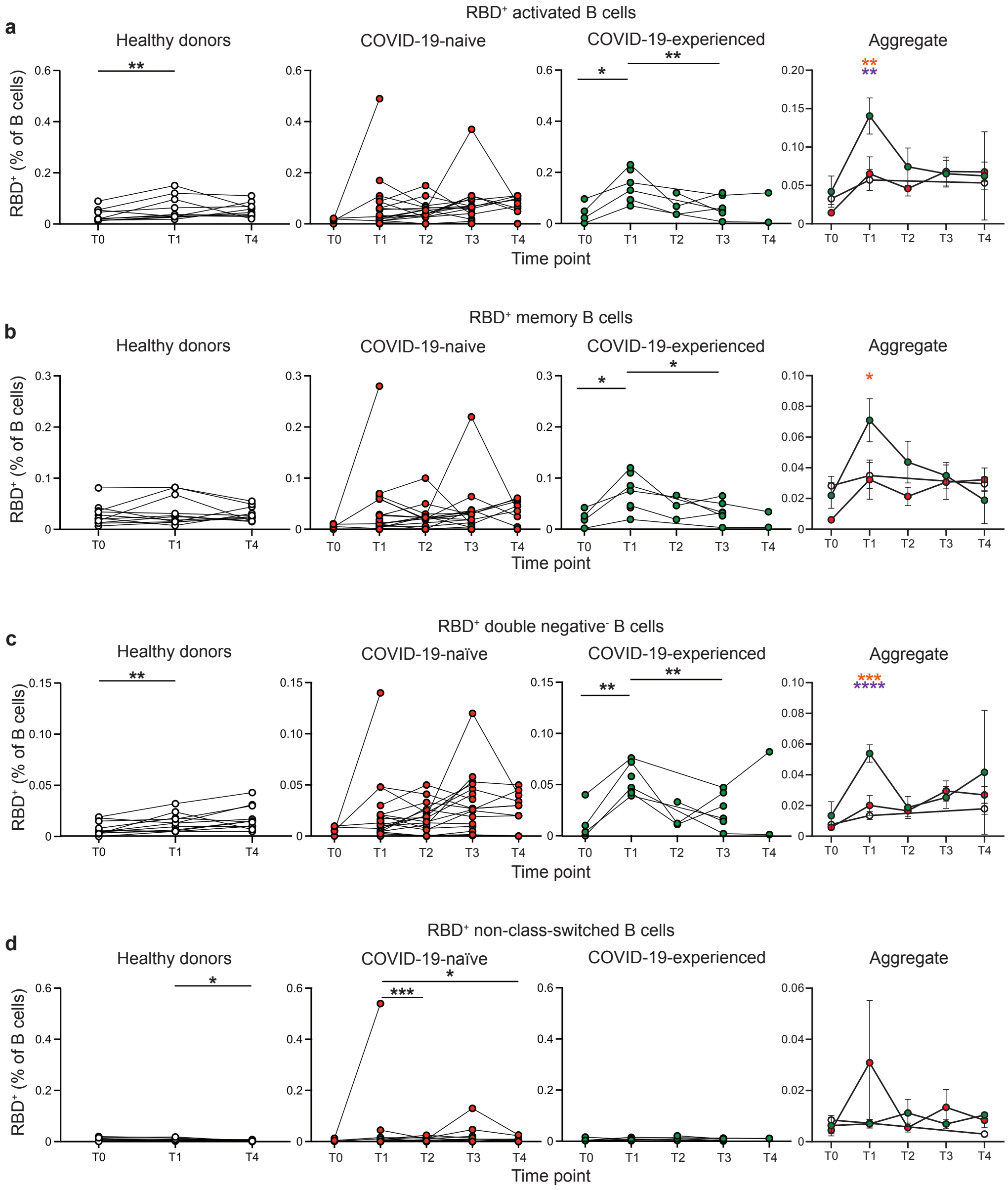

RBD⁺ activated B cells
a
Healthy donors
COVID-19-naive
COVID-19-experienced
Aggregate
RBD⁺ (% of B cells)
Time point
RBD⁺ memory B cells
b
RBD⁺ double negative⁻ B cells
c
COVID-19-naïve
RBD⁺ non-class-switched B cells
d
T0
T1
T2
T3
T4


**Figure S2**

**Figure S2**. **COVID-19-experienced PAD patients display elevated RBD-specific B cell response following primary vaccination series**. (**a**) Percentage of activated RBD$^+$ cells amongst the B (Live CD19$^+$ CD3$^-$) cell population in the healthy donor (left, white), COVID-19-naïve PAD (middle left, red), and COVID-19-experienced PAD (middle right, green) cohorts. Aggregate of mean percentage of B cells that are RBD$^+$ activated cells in all groups is shown on right. (**b**) Percentage of memory (IgD$^{lo}$ CD20$^+$ CD38$^{int\text{-}lo}$ CD27$^+$) RBD$^+$ cells amongst the B cell population in the healthy donor (left, white), COVID-19-naïve PAD (middle, red), and COVID-19-experienced PAD (middle, green) cohorts. Aggregate of mean percentage of B cells that are RBD$^+$ memory cells in all groups is shown on right. (**c**) Percentage of double negative (IgD$^{lo}$ CD20$^+$ CD38$^{int\text{-}lo}$ CD27$^-$) RBD$^+$ cells amongst the B cell population in the healthy donor (left, white), COVID-19-naïve PAD (middle, red), and COVID-19-experienced PAD (middle, green) cohorts. Aggregate of mean percentage of B cells that are double negative RBD$^+$ cells in all groups is shown on right. (**d**) Percentage of non-class-switched (IgD$^+$ CD20$^+$ CD38$^{int\text{-}lo}$ CD27$^+$) RBD$^+$ cells amongst the B cell population in the healthy donor (left, white), COVID-19-naïve PAD (middle, red), and COVID-19-experienced PAD (middle, green) cohorts. Aggregate of mean percentage of B cells that are RBD$^+$ non-class-switched cells in all groups is shown on right. Statistical analyses were performed using a mixed effects model (for trends found between time points) or two-way ANOVA (for trends found between groups in the aggregate graphs) with Fisher's least significant difference testing. Significance testing between time points was limited to comparisons relative to T1. On the aggregate graphs, all error bars were calculated based on the standard error of the mean. Above the aggregate graphs, an orange asterisk indicates a comparison between the COVID-19-naïve and COVID-19-experienced groups, and a purple asterisk indicates a comparison between the COVID-19-experienced and healthy donor groups (*, $p < 0.05$; **, $p < 0.01$; ***, $p<0.001$; ****, $p<0.001$).

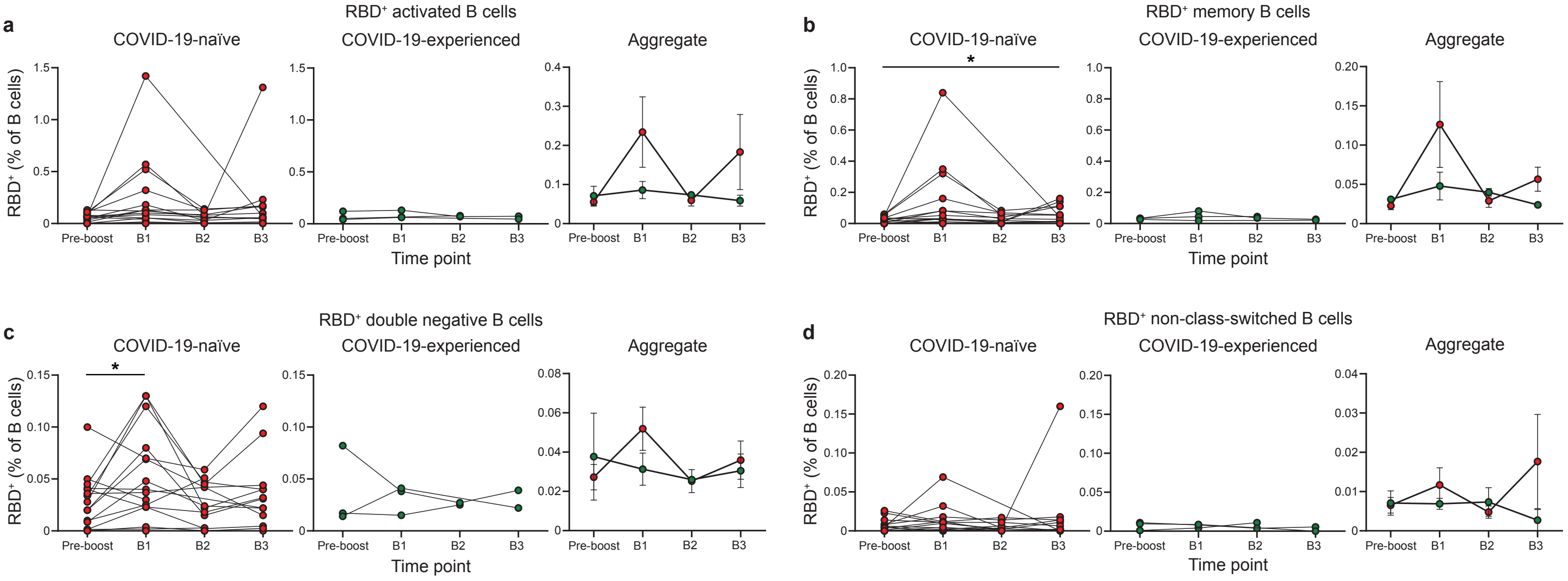

a
RBD⁺ activated B cells
COVID-19-naïve
COVID-19-experienced
Aggregate
RBD⁺ (% of B cells)
Pre-boost
B1
B2
B3
Time point
b
RBD⁺ memory B cells
c
RBD⁺ double negative B cells
d
RBD⁺ non-class-switched B cells


**Figure S3**

**Figure S3. COVID-19-naive PAD patients display elevated RBD-specific B cell response following booster vaccination.** (**a**) Percentage of activated RBD$^+$ cells amongst the B (Live CD19$^+$ CD3$^-$) cell population in the healthy donor (left, white), COVID-19-naïve PAD (middle left, red), and COVID-19-experienced PAD (middle right, green) cohorts. Aggregate of mean percentage of B cells that are RBD$^+$ activated cells in all groups is shown on right. (**b**) Percentage of memory (IgD$^{lo}$ CD20$^+$ CD38$^{int\text{-}lo}$ CD27$^+$) RBD$^+$ cells amongst the B cell population in the healthy donor (left, white), COVID-19-naïve PAD (middle, red), and COVID-19-experienced PAD (middle, green) cohorts. Aggregate of mean percentage of B cells that are RBD$^+$ memory cells in all groups is shown on right. (c) Percentage of double negative (IgD$^{lo}$ CD20$^+$ CD38$^{int\text{-}lo}$ CD27$^-$) RBD$^+$ cells amongst the B cell population in the healthy donor (left, white), COVID-19-naïve PAD (middle, red), and COVID-19-experienced PAD (middle, green) cohorts. Aggregate of mean percentage of B cells that are double negative Spike$^+$ cells in all groups is shown on right. (**d**) Percentage of non-class-switched (IgD$^+$ CD20$^+$ CD38$^{int\text{-}lo}$ CD27$^+$) RBD$^+$ cells amongst the B cell population in the healthy donor (left, white), COVID-19-naïve PAD (middle, red), and COVID-19-experienced PAD (middle, green) cohorts. Aggregate of mean percentage of B cells that are RBD$^+$ non-class-switched cells in all groups is shown on right. pre-boost group consists of the last sample obtained from each patient prior to booster vaccination. Statistical analyses were performed using a mixed effects model (for trends found between time points) with Fisher's least significant difference testing. Significance testing between time points was limited to comparisons relative to pre-boost (*, $p < 0.05$). On the aggregate graphs, error bars were calculated based on the standard error of the mean.

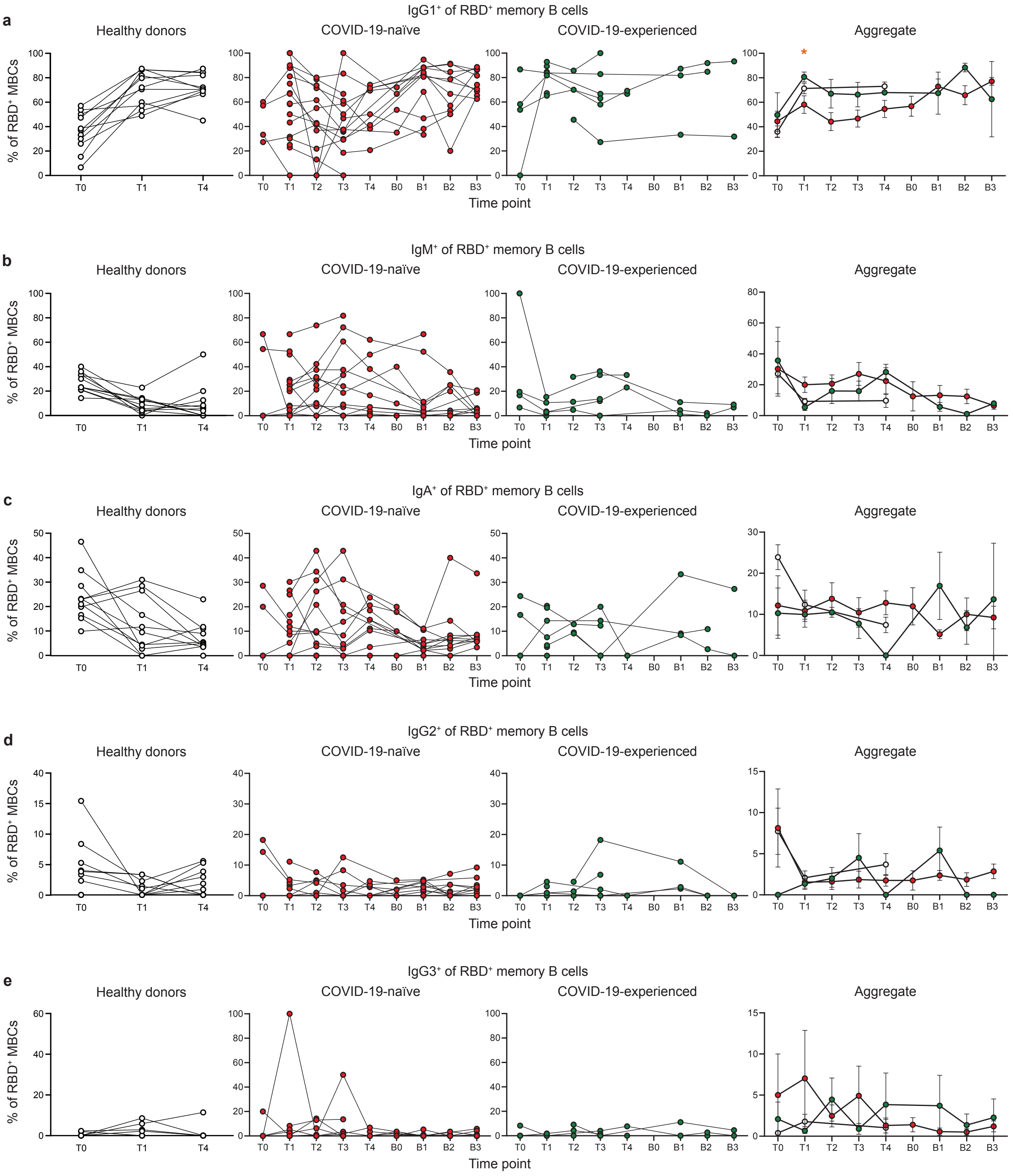

a
IgG1⁺ of RBD⁺ memory B cells
b
IgM⁺ of RBD⁺ memory B cells
c
IgA⁺ of RBD⁺ memory B cells
d
IgG2⁺ of RBD⁺ memory B cells
e
IgG3⁺ of RBD⁺ memory B cells
Healthy donors
COVID-19-naïve
COVID-19-experienced
Aggregate
% of RBD⁺ MBCs
Time point
T0
T1
T2
T3
T4
B0
B1
B2
B3
*


**Figure S4**

**Figure S4. Isotype composition of SARS-CoV-2 RBD-specific memory B cell response following vaccination in PAD patients**. (**a**) Percentage of RBD$^+$ memory B cells that are IgG1$^+$ in the healthy donor (left, white), COVID-19-naïve PAD (middle left, red), and COVID-19-experienced PAD (middle right, green) cohorts. Aggregate of mean percentage of IgG1$^+$ cells in all groups is shown on right. (**b**) Percentage of RBD$^+$ memory B cells that are IgM$^+$ in the healthy donor (left, white), COVID-19-naïve PAD (middle left, red), and COVID-19-experienced PAD (middle right, green) cohorts. Aggregate of mean percentage of IgM$^+$ cells in all groups is shown on right. (**c**) Percentage of RBD$^+$ memory B cells that are IgA$^+$ in the healthy donor (left, white), COVID-19-naïve PAD (middle left, red), and COVID-19-experienced PAD (middle right, green) cohorts. Aggregate of mean percentage of IgA$^+$ cells in all groups is shown on right. (**d**) Percentage of RBD$^+$ memory B cells that are IgG2$^+$ in the healthy donor (left, white), COVID-19-naïve PAD (middle left, red), and COVID-19-experienced PAD (middle right, green) cohorts. Aggregate of mean percentage of IgG2$^+$ cells in all groups is shown on right. (**e**) Percentage of RBD$^+$ memory B cells that are IgG3$^+$ in the healthy donor (left, white), COVID-19-naïve PAD (middle left, red), and COVID-19-experienced PAD (middle right, green) cohorts. Aggregate of mean percentage of IgG3$^+$ cells in all groups is shown on right. Statistical analyses were performed using a two-way ANOVA (for trends found between groups in the aggregate graphs) with Fisher's least significant difference testing. On the aggregate graphs, error bars were calculated based on the standard error of the mean. Above the aggregate graphs, an orange asterisk indicates a comparison between the COVID-19-naïve and COVID-19-experienced groups (*, $p < 0.05$).

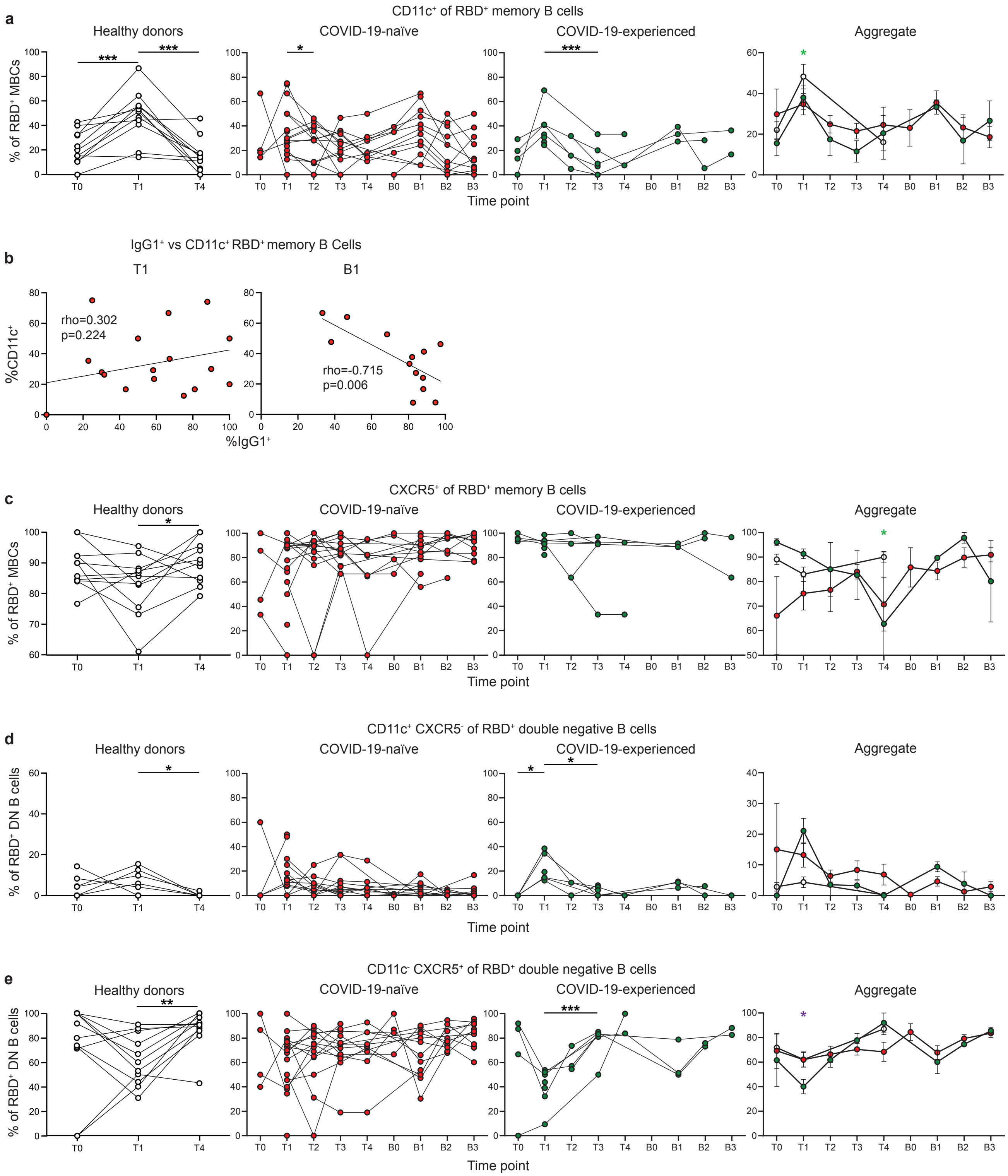

a
CD11c⁺ of RBD⁺ memory B cells
Healthy donors
COVID-19-naïve
COVID-19-experienced
Aggregate
% of RBD⁺ MBCs
Time point
b
IgG1⁺ vs CD11c⁺ RBD⁺ memory B Cells
T1
B1
%CD11c⁺
%IgG1⁺
rho=0.302
p=0.224
rho=-0.715
p=0.006
c
CXCR5⁺ of RBD⁺ memory B cells
Healthy donors
COVID-19-naïve
COVID-19-experienced
Aggregate
% of RBD⁺ MBCs
Time point
d
CD11c⁺ CXCR5⁻ of RBD⁺ double negative B cells
Healthy donors
COVID-19-naïve
COVID-19-experienced
Aggregate
% of RBD⁺ DN B cells
Time point
e
CD11c⁻ CXCR5⁺ of RBD⁺ double negative B cells
Healthy donors
COVID-19-naïve
COVID-19-experienced
Aggregate
% of RBD⁺ DN B cells
Time point


**Figure S5**

**Figure S5. RBD-specific memory B cells from PAD patients display reduced CD11c expression class**. (**a**) Percentage of $RBD^+$ memory B cells that are $CD11c^+$ in the healthy donor (left, white), COVID-19-naïve PAD (middle left, red), and COVID-19-experienced PAD (middle right, green) cohorts. Aggregate of mean percentage of $CD11c^+$ cells in all groups is shown on right. (**b**) Correlation between percentage of $RBD^+$ memory B cells that are $IgG1^+$ and $CD11c^+$ at T1 (left) or B1 (right). Associations for **b** are calculated using Pearson rank correlation and are shown with Pearson trend lines for visualization. (**c**) Percentage $RBD^+$ memory B cells that are $CXCR5^+$ in the healthy donor (left, white), COVID-19-naïve PAD (middle left, red), and COVID-19-experienced PAD (middle right, green) cohorts. Aggregate of mean percentage of $CXCR5^+$ cells in all groups is shown on right. (**d**) Percentage of $RBD^+$ double negative B cells that are $CD11c^+$ $CXCR5^-$ in the healthy donor (left, white), COVID-19-naïve PAD (middle left, red), and COVID-19-experienced PAD (middle right, green) cohorts. Aggregate of mean percentage of are $CD11c^+$ $CXCR5^-$ cells in all groups is shown on right. (**e**) Percentage of $RBD^+$ double negative B cells that are $CD11c^-$ $CXCR5^+$ in the healthy donor (left, white), COVID-19-naïve PAD (middle left, red), and COVID-19-experienced PAD (middle right, green) cohorts. Aggregate of mean percentage of are $CD11c^-$ $CXCR5^+$ cells in all groups is shown on right. Statistical analyses in a, c-e were performed using mixed effects model (for trends found between time points) or a two-way ANOVA (for trends found between groups in the aggregate graphs) with Fisher's least significant difference testing. Significance testing between time points was limited to comparisons relative to T1. On the aggregate graphs, error bars were calculated based on the standard error of the mean. Above the aggregate graphs, a green asterisk indicates a comparison between the COVID-19-naïve and healthy donor groups, a purple asterisk indicates a comparison between the COVID-19-experienced and healthy donor groups (*, $p < 0.05$; **, $p < 0.01$; ***, $p < 0.001$).

| Patient Number | Age range | Sex | Vaccine Type | Booster Type | Diagnosis | COVID-19 infection to 1st vaccine (days) |
|---|---|---|---|---|---|---|
| 101 | 56-60 | F | Pfizer | Pfizer | CVID | -- |
| 102 | 21-25 | F | Pfizer | Pfizer | CVID | -- |
| 103 | 56-60 | F | Pfizer | -- | CVID | -- |
| 104 | 41-45 | F | Pfizer | Pfizer | CVID | -- |
| 105 | 26-30 | M | Pfizer | Pfizer | CVID | -- |
| 106 | 61-65 | F | Pfizer | -- | CVID | -- |
| 107 | 71-75 | F | Pfizer | Pfizer | SAB | 96 |
| 108 | 61-65 | F | Pfizer | Pfizer | CVID | -- |
| 109 | 36-40 | F | J&J | Pfizer | SAB | -- |
| 110 | 46-50 | F | Pfizer | Pfizer | CVID | -- |
| 111 | 56-60 | F | Pfizer | Pfizer | CVID | -- |
| 112 | 41-45 | M | Moderna | Moderna | CVID | -- |
| 114 | 31-35 | F | Pfizer | -- | SAB | 90 |
| 115 | 16-20 | F | Moderna | -- | CVID | 181 |
| 116 | 26-30 | F | J&J | -- | SAB | -- |
| 117 | 81-85 | F | Moderna | -- | CVID | -- |
| 118 | 61-65 | F | Moderna | -- | CVID | -- |
| 119 | 21-25 | F | Pfizer | Pfizer | Hypogam | 276 |
| 120 | 41-45 | F | Moderna | Moderna | CVID | -- |
| 121 | 66-70 | F | Pfizer | Pfizer | SAB | -- |
| 122 | 46-50 | F | Pfizer | -- | CVID | 117 |
| 123 | 66-70 | F | Pfizer | Pfizer | Hypogam | -- |
| 124 | 51-55 | F | Moderna | -- | CVID | 222 |
| 125 | 56-60 | F | Moderna | -- | Hypogam | 106 |
| 126 | 56-60 | M | J&J | Pfizer | CVID | -- |
| 127 | 56-60 | F | Pfizer | Pfizer | CVID | -- |
| 128 | 61-65 | F | Pfizer | Pfizer | Hypogam | -- |
| 129 | 36-40 | F | Pfizer | Pfizer | CVID | -- |
| 130 | 46-50 | F | Moderna | Moderna | SAB | 144 |
| 131 | 26-30 | F | Pfizer | -- | CVID | 36 |
| 368-05 | 36-40 | M | Pfizer | -- | Healthy donor | -- |
| 368-17 | 36-40 | M | Pfizer | -- | Healthy donor | -- |
| 368-24 | 51-55 | F | Pfizer | -- | Healthy donor | -- |
| 368-25 | 41-45 | F | Pfizer | -- | Healthy donor | -- |
| 368-27 | 46-50 | M | Pfizer | -- | Healthy donor | -- |
| 368-29 | 26-30 | F | Pfizer | -- | Healthy donor | -- |
| 368-34 | 26-30 | M | Pfizer | -- | Healthy donor | -- |
| 368-36 | 46-50 | F | Pfizer | -- | Healthy donor | -- |
| 368-37 | 41-45 | M | Pfizer | -- | Healthy donor | -- |
| 368-38 | 41-45 | F | Pfizer | -- | Healthy donor | -- |
| 368-40 | 31-35 | F | Pfizer | -- | Healthy donor | -- |

**Table S1. Characteristics of patient cohort**

F, female; M, male; CVID, common variable immune defeiciency; hypogam, hypogammaglobulinemia; SAB, specific antibody deficiency disorder.

| Patient Number | B cells (% of Live) | IgD$^{lo}$ (% of CD19$^{+}$) | IgD$^{lo}$ CD27$^{+}$ (% of CD19$^{+}$) | CD3$^{+}$ T cells (% of Live) | CD4$^{+}$ T cells (% of CD3$^{+}$) | CD8$^{+}$ T cells (% of CD3$^{+}$) | CD4$^{+}$/CD8$^{+}$ Ratio |
|---|---|---|---|---|---|---|---|
| 101 | 4.07 | 34.3 | 18.8 | 34.6 | 57.6 | 10 | 5.76 |
| 102 | 5.21 | 12.5 | 5.16 | 28.3 | 47.4 | 28.2 | 1.68 |
| 103 | 4.74 | 11.7 | 4.61 | 50.2 | 51.4 | 24.5 | 2.10 |
| 104* | 2.9 | 7.81 | 0.53 | 7.72 | 44.2 | 13.2 | 3.35 |
| 105 | 4.21 | 11.4 | 5.51 | 10.3 | 32.4 | 26.1 | 1.24 |
| 106* | 6.13 | 5.63 | 1.68 | 12.7 | 52.9 | 8.48 | 6.24 |
| 107 | 3.02 | 27.1 | 15.1 | 17.1 | 42.9 | 18.9 | 2.27 |
| 108 | 4.42 | 30.6 | 17.1 | 11.8 | 45.7 | 15.1 | 3.03 |
| 109 | 9.89 | 12.8 | 8.27 | 16.5 | 48.6 | 16.9 | 2.88 |
| 110 | 3.61 | 32 | 18.1 | 18.1 | 49.3 | 18 | 2.74 |
| 111 | 4.01 | 44.5 | 34.5 | 15.2 | 47.8 | 16 | 2.99 |
| 112* | 8.5 | 10.7 | 1.53 | 14.9 | 34.6 | 36.5 | 0.95 |
| 114 | 6.65 | 30.9 | 20.1 | 24.9 | 56.4 | 16.3 | 3.46 |
| 115 | 6.78 | 24.6 | 18.1 | 15 | 43.6 | 22.9 | 1.90 |
| 116 | 1.61 | 18.8 | 11.9 | 15.7 | 38.1 | 27.3 | 1.40 |
| 117 | 4.45 | 14.5 | 4.97 | 17.2 | 50.1 | 4.8 | 10.4 |
| 118 | 4.67 | 14.7 | 10.3 | 17.6 | 46.5 | 15.6 | 2.98 |
| 119 | 4.95 | 22.8 | 11.9 | 15.3 | 47.5 | 17.4 | 2.73 |
| 120 | 6.99 | 31.7 | 11.9 | 17.6 | 46.1 | 11.3 | 4.08 |
| 121 | 7.43 | 21.8 | 18.2 | 19.6 | 54.9 | 8.01 | 6.85 |
| 122 | 6.54 | 48.9 | 11 | 18.6 | 39.9 | 9.3 | 4.29 |
| 123 | 8.75 | 10.9 | 39.8 | 17.6 | 55 | 5.18 | 10.6 |
| 124 | 8.08 | 17.9 | 4.09 | 17.2 | 48.6 | 21.5 | 2.26 |
| 125 | 10.1 | 13.3 | 11.3 | 19.9 | 50.3 | 21.5 | 2.34 |
| 126* | 5.89 | 11.4 | 6.62 | 17.7 | 48.5 | 14 | 3.46 |
| 127 | 4.92 | 17.1 | 4.82 | 19.6 | 56.7 | 16.4 | 3.46 |
| 128 | 2.77 | 29.5 | 3.09 | 16.3 | 44.9 | 19.9 | 2.26 |
| 129 | 8.45 | 8.89 | 20.8 | 17.5 | 44.1 | 26.6 | 1.66 |
| 130 | 6.74 | 33.9 | 3.54 | 18.4 | 46 | 27.2 | 1.69 |
| 131 | 4.43 | 8.39 | 25.3 | 12.7 | 39.1 | 31.2 | 1.25 |
| 368-05 | 10.7 | 12.3 | 8.52 | 30.8 | 47.3 | 32 | 1.48 |
| 368-17 | 8.85 | 25.1 | 15.1 | 19.9 | 57.5 | 8.68 | 6.62 |
| 368-24 | 7.82 | 16.3 | 11.8 | 28.7 | 60.8 | 12.1 | 5.02 |
| 368-25 | 13.1 | 22.9 | 17.3 | 21 | 48.9 | 10.4 | 4.70 |
| 368-27 | 3.12 | 24.8 | 22.4 | 44.5 | 33.3 | 22.3 | 1.49 |
| 368-29 | 8.19 | 21.3 | 14.1 | 26.6 | 48.3 | 19.2 | 2.52 |
| 368-34 | 15.7 | 8.93 | 6.25 | 33.6 | 43.7 | 11.3 | 3.87 |
| 368-36 | 6.64 | 20.8 | 16 | 24.2 | 54 | 16.3 | 3.31 |
| 368-37 | 6.88 | 23.9 | 18.5 | 22.6 | 48.4 | 6.98 | 6.93 |
| 368-38 | 7.05 | 7.13 | 4.5 | 11.7 | 44.7 | 10.1 | 4.43 |
| 368-40 | 9.09 | 27.1 | 17.6 | 24.3 | 48.2 | 16 | 3.01 |

**Table S2. Cellular phenotype of patient cohort**

* Patients classified as non-responders (IgDlo Spike+ B cell response <0.02% of total B cells at day 7 to 28 post vaccination).

| Patient number | Most Recent CBC | | Lymphocyte Subpopulation at the Time of Diagnosis | | | | | Immunoclobulin levels at the Time of Diagnosis | | | Most Recent IgG |
|---|---|---|---|---|---|---|---|---|---|---|---|
| | WBC | ALC | CD3 | CD4 | CD8 | CD19 | CD16/56 | IgG | IgA | IgM | |
| **Normal range** | **3.8 - 9.9 (1000s per mm$^3$)** | **1,000 - 3,300 (per mm$^3$)** | **661 - 1963 (per mm$^3$)** | **490 - 1294 (per mm$^3$)** | **187 - 781 (per mm$^3$)** | **110 - 488 (per mm$^3$)** | **76 - 467 (per mm$^3$)** | **700 - 1600 mg/dL** | **75 - 400 mg/dL** | **40 - 230 mg/dL** | **mg/dL** |
| 101 | 5.8 | 2400 | 2118 | 1668 | 470 | 283 | 258 | 477 | 53 | 62 | 1065 |
| 102 | 4 | 1000 | 895 | 426 | 432 | 227 | 127 | 560 | 62 | 24 | 1134 |
| 103 | 4.5 | 1674 | 1450 | 934 | 456 | 187 | 180 | 566 | 140 | 106 | 828 |
| 104* | 5.1 | 1000 | 818 | 601 | 193 | 251 | 18 | <40 | <4 | <5 | 725 |
| 105 | 12 | 1500 | 974 | 384 | 487 | 280 | 221 | <300 | <10 | <25 | 947 |
| 106* | 7.2 | 1600 | -- | -- | -- | -- | -- | 432 | 89 | 95 | 830 |
| 107 | 6.5 | 1900 | 1109 | 775 | 294 | 80 | 88 | 514 | 72 | 165 | 505 |
| 108 | 9.5 | 3500 | Reported normal in immunology clinic note | | | | | 529 | 188 | 35 | 938 |
| 109 | 6.4 | 1100 | -- | -- | -- | -- | -- | 895 | 142 | 105 | 1345 |
| 110 | 4.8 | 1700 | 1632 | 859 | 738 | 184 | 59 | 1304 | 82 | 37 | 1462 |
| 111 | 5.2 | 860 | -- | -- | -- | -- | -- | 465 | -- | -- | 836 |
| 112* | 4.3 | 1000 | 1162 | 375 | 663 | 134 | 151 | 1850 | <25 | <19 | 1303 |
| 114 | 8.3 | 1900 | 1702 | 1054 | 547 | 243 | 41 | 911 | 221 | 58 | 1291 |
| 115 | 15.6 | 2400 | -- | -- | -- | -- | -- | 340 | 12 | reported nl | 1261 |
| 116 | 8 | 2400 | 1974 | 1167 | 690 | 262 | 71 | 983 | 162 | 52 | 983 |
| 117 | 5.4 | 1600 | 2194 | 1646 | 549 | 462 | 144 | 553 | 175 | 49 | 1143 |
| 118 | 7.3 | 1700 | Reported normal in immunology clinic note | | | | | 647 | <4 | <5 | 758 |
| 119 | 5 | 2200 | 2055 | 1396 | 790 | 211 | 79 | 473 | 120 | 167 | 877 |
| 120 | 8.3 | 2600 | 1848 | 1282 | 501 | 157 | 199 | 528 | 136 | 155 | 1164 |
| 121 | 6.2 | 2400 | 859 | 716 | 119 | 95 | 227 | 622 | 133 | 137 | 989 |
| 122 | 10.6 | 2720 | 3300 | 2274 | 1026 | 580 | 446 | 606 | 204 | 30 | 743 |
| 123 | 8 | 1500 | 1055 | 835 | 205 | 315 | 158 | 665 | 162 | 45 | 887 |
| 124 | 9.1 | 1190 | Reported low NK cells only in immunology clinic note | | | | | 502 | 42 | 95 | 547 |
| 125 | 4.9 | 2000 | 1837 | 1086 | 725 | 475 | 308 | 656 | 270 | 10 | 657 |
| 126* | 4.5 | 1600 | Reported low CD19 at 39, all others normal in immunology note | | | | | <200 | 66 | <25 | 860 |
| 127 | 3.8 | 1400 | 925 | 638 | 287 | 64 | 43 | 516 | <25 | 521 | 781 |
| 128 | 7.2 | 800 | 655 | 389 | 236 | 209 | 78 | 629 | 196 | 80 | 990 |
| 129 | 5 | 1420 | -- | -- | -- | -- | -- | 261 | <10 | <25 | 1240 |
| 130 | 6.27 | 1610 | -- | -- | -- | -- | -- | 814 | 99 | 198 | 1145 |
| 131 | 10.79 | 1700 | -- | -- | -- | -- | -- | 346 | <10 | <25 | 1036 |

**Table S3. PAD patient laboratory values**
CBC - complete blood count; WBC - white blood count, ALC - absolute lymphocyte count; Ig - immunoglobulin; dL - deciliter. Empty cells (dashed lines) indicate that the test was not performed.

* Patients classified as non-responders (IgDlo Spike+ B cell response <0.02% of total B cells at day 7 to 28 post vaccination).

| Patient | *S. pneumoniae* Titer** | Tetanus Titer IU/ml**** | Diphtheria Titer IU/ml**** |
|---|---|---|---|
| 101 | 8/23 | N/A | -- |
| 102 | 7/23 | 0.51 | -- |
| 103 | 10/23 | Positive | Positive |
| 104* | 0/23 | Positive | -- |
| 105 | 0/23*** | Negative | -- |
| 106* | 4/13 | 4.59 | -- |
| 107 | 14/23 | 0.97 | 0.09 |
| 108 | 2/23 | 0.06 | 0.01 |
| 109 | 6/23 | >7 | 2.34 |
| 110 | 5/23 | N/A | -- |
| 111 | 9/14 | 0.98 | -- |
| 112* | --- | N/A | -- |
| 114 | 12/23 | 1.56 | -- |
| 115 | Unprotective | -- | -- |
| 116 | 14/23 | 0.36 | Positive |
| 117 | 11/23 | 0.01 | -- |
| 118 | -- | -- | -- |
| 119 | 19/23 | 0.45 | -- |
| 120 | 14/23 | >2.24 | -- |
| 121 | 8/23 | 0.46 | -- |
| 122 | 10/23 | 0.72 | -- |
| 123 | 20/23 | >2.24 | -- |
| 124 | 2/24 | -- | -- |
| 125 | 7/23 | 2.2 | -- |
| 126* | 3/24*** | 0.24 | Undetectable |
| 127 | 0/13 | -- | -- |
| 128 | 14/23 | 0.87 | 0.07 |
| 129 | Unprotective | -- | -- |
| 130 | 0/23 | 2.75 | -- |
| 131 | -- | 1.73 | 0.35 |

**Table S4. PAD patient responses to other vaccine antigens at the time of diagnosis**

IU - international unit. N/A - data was obtained but not available in the medical record. Dashed lines indicate that the test was not performed.

* Patients classified as non-responders (IgDlo Spike+ B cell response <0.02% of total B cells at day 7 to 28 post vaccination).

** Indicates the number of tested anti-*Streptococcus pneumoniae* serotypes with a level above 1.3 μg/ml for ≥17 tested serotypes.

*** Pre-pneumovax booster values; patient started on immunoglobulin replacement prior to post-booster recheck.

**** A value of ≥ 0.01 IU/ml is considered positive. In some records, a positive notation was indicated but no value was provided.
